# Impact of bivalent BA.4/5 BNT162b2 COVID-19 vaccine on acute symptoms, quality of life, work productivity and activity levels among symptomatic US adults testing positive for SARS-CoV-2 at a national retail pharmacy

**DOI:** 10.1101/2023.09.21.23295904

**Authors:** Manuela Di Fusco, Xiaowu Sun, Laura Anatale-Tardiff, Alon Yehoshua, Henriette Coetzer, Mary B. Alvarez, Kristen E. Allen, Thomas M. Porter, Laura Puzniak, Santiago M.C Lopez, Joseph C. Cappelleri

## Abstract

**Background:** Evidence on the impact of COVID-19 vaccination on symptoms, Health-Related Quality of Life (HRQoL), Work Productivity and Activity Impairment (WPAI) is scarce. We analyzed associations between bivalent BA.4/5 BNT162b2 and these patient-reported outcomes (PROs).

**Methods:** Symptomatic US adults who tested positive for SARS-CoV-2 were recruited between 03/02-05/18/2023. PROs were assessed using a CDC-based symptom questionnaire, EQ-5D-5L, WPAI-GH, and PROMIS Fatigue, from pre-COVID to Week 4 following infection. Multivariable analysis using mixed models for repeated measures was conducted, adjusting for several covariates.

**Results:** The study included 641 participants: 314 vaccinated with bivalent BA.4/5 BNT162b2 and 327 unvaccinated/not up-to-date. Mean (SD) age was 46.5 years (15.9), 71.2% were female, 44.2% reported prior infection, 25.7% had ≥1 comorbidity. The BA.4/5 BNT162b2 cohort reported fewer acute symptoms through Week 4, especially systemic and respiratory symptoms. All PROs were adversely affected, especially at Week 1; however, at that time point, the bivalent BA.4/5 BNT162b2 cohort reported better work performance, driven by less absenteeism, and fewer work hours lost. No significant differences were observed for HRQoL.

**Conclusions:** COVID-19 negatively impacted patient outcomes. Compared with unvaccinated/not up-to-date participants, those vaccinated with bivalent BA.4/5 BNT162b2 reported fewer and less persistent symptoms and improved work performance.

Clinicaltrials.gov NCT05160636

## BACKGROUND

A growing body of evidence indicates that COVID-19 has profound implications on patients’ wellbeing and social function [1,2]. The multiorgan symptoms of SARS-CoV-2 infection have been associated with a decline in health-related quality of life (HRQoL), impairments in daily activities, and the ability to work [3–5].

From the original monovalent vaccines to the bivalent vaccines, COVID-19 vaccination significantly impacted the global COVID-19 response. Evidence on the efficacy, safety and effectiveness of mRNA COVID-19 vaccination is extensive for the original monovalent formulation and is rapidly growing for the bivalent formulation as well[6–8]. The mRNA bivalent vaccines targeted both the original strain and BA.4/BA.5 Omicron lineages and were authorized for use as a single booster on 8/31/2022 to provide further protection against Omicron, replacing the original monovalent formulations[9].

The BNT162b2 COVID-19 vaccine has shown additive benefits beyond traditional endpoints [3–5]. In a previous nationwide study of symptomatic US outpatients, being boosted with the original monovalent BNT162b2 was associated with a significant reduction in the prevalence and persistence of both acute and long-term symptoms, and improved health-related quality of life, activity levels and work performance[4,5]. There is a dearth of such data for the bivalent formulation. This study sought to address these gaps and assess the burden of COVID-19 infection and the impact of the Pfizer-BioNTech COVID-19 vaccine (original and BA.4/5 bivalent) on symptoms, HRQoL, work productivity and activity impairment prior to through one month following SARS-CoV-2 infection.

## METHODS

### Study Design and Participants

This was a nationwide prospective patient-reported outcomes (PRO) survey-based study that leveraged a previously described design (clinicaltrials.gov NCT05160636) [3–5]. The source population consisted of individuals testing for SARS-CoV-2 at one of approximately 5,000 CVS Health test sites across the United States (US). As part of the registration process for scheduling a SARS-CoV-2 test at a CVS Health site, individuals are required to complete a screening questionnaire including demographics, symptoms, comorbidities, and COVID-19 vaccination status. The screening variables, reverse transcription–polymerase chain reaction (RT-PCR) test and rapid antigen test results are loaded in an analytic dataset, where ∼80-90% of test results are reported within 2-3 days from the date of the test appointment. Leveraging this analytic platform, this study was designed as a prospective survey-based patient-reported outcomes study targeting adults 18 years of age and older with a positive RT-PCR or rapid antigen test result and self-reporting at least one acute COVID-19 symptom. The study excluded asymptomatic patients. The individuals meeting the inclusion and exclusion criteria were emailed an invitation as soon as their test results became available, no later than 4 days from testing and were recruited between 03/02/2023 and 05/18/2023 during the predominance of XBB Omicron sub-lineage circulation (Supplemental Figure 1). The email invitation directed the potential participants to a dedicated e-consent website to learn about the study, the survey schedule and to sign an informed consent. To encourage participation, email reminders were sent throughout the study follow-up period, and the survey was voluntary and anonymous. To minimize data missingness, respondents could not skip any surveys. Participants could discontinue participation from the research study at any time.

### Study Cohorts

At enrollment, we categorized the study participants based on their pre-infection COVID-19 vaccination history. The study population for these analyses included two mutually exclusive cohorts: a “Bivalent” and an “Unvaccinated” cohort (Figure 1). Participants were included in the bivalent cohort if in the pre-test screening questionnaire they reported a date of 9/1/22 or later[9] for their most recent dose of the Pfizer-BioNTech COVID-19 vaccine (original and BA.4/5 bivalent) COVID-19 Vaccine. From this date, the bivalent was the only formulation available and authorized in the US [9]. Participants were considered unvaccinated if they did not report any COVID-19 vaccine prior to testing or if they reported being primed (fully vaccinated) with their last monovalent dose received over 12 months before enrollment, the time at which their vaccine-induced immunity was assumed to have waned completely[10]. As such, this cohort comprised both unvaccinated and not up-to-date participants. For simplicity, this cohort is defined as “Unvaccinated” and this term is used throughout this report. To confirm vaccination status, participants’ subsequent responses to vaccination date questions were compared with their index responses (at time of registering for testing); if responses did not match, the information was queried and adjudicated, and the latest information was used.

**Figure 1.**
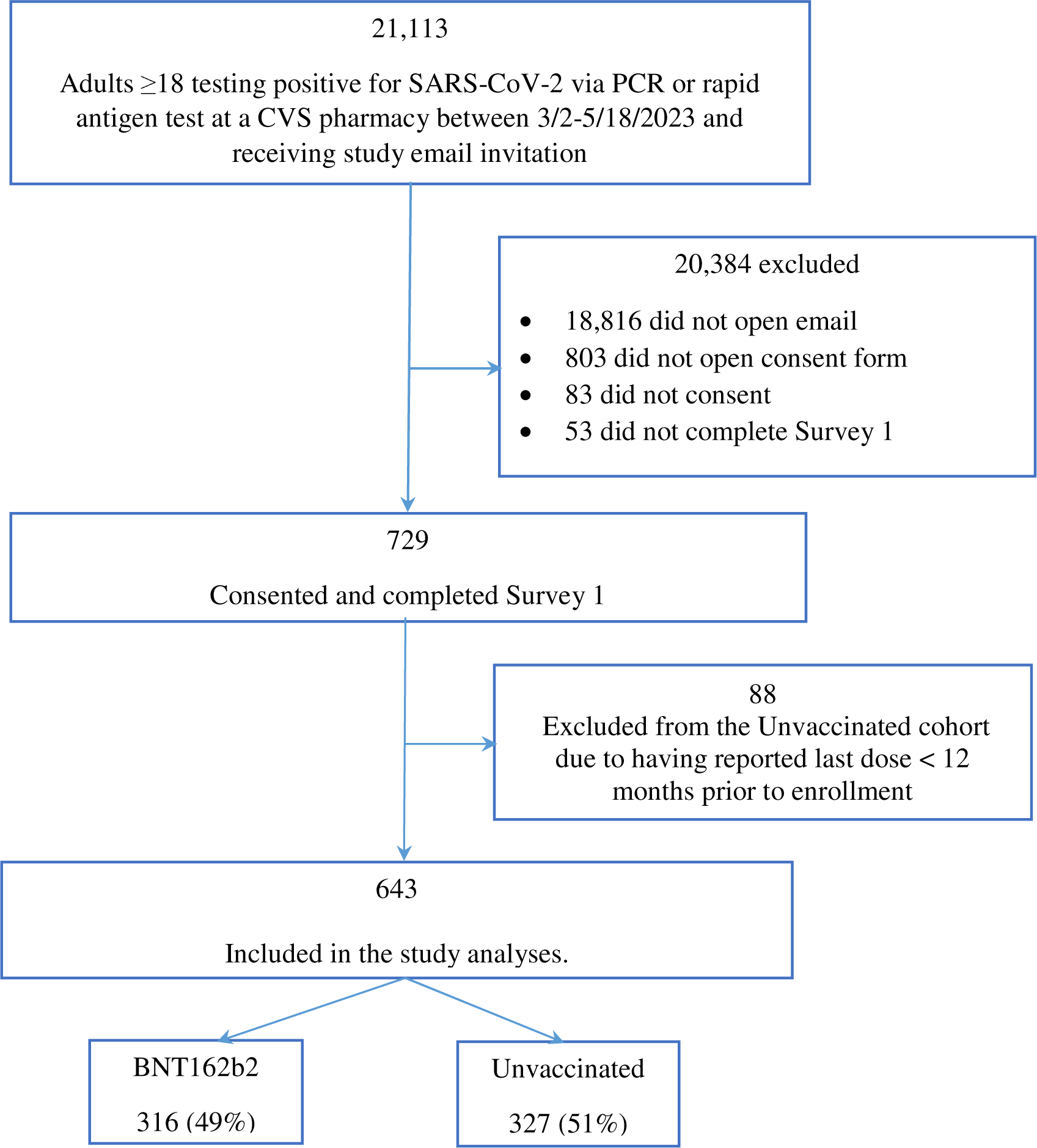
Study participant flowchart.

### Baseline Characteristics and Patient Reported Outcome Measures

The baseline characteristics of study participants were obtained via the CVS Health pre-test screening questionnaire, which comprised demographics, comorbidities, COVID-19 vaccination and COVID-19 infection history. The Social Vulnerability Index (SVI) was calculated based on zip codes, with a value of 0 representing the lowest level of vulnerability and a value of 1 representing highest vulnerability [11]. An additional questionnaire captured COVID-19 antiviral treatment use and changes in vaccination and infection status at each study time point. The study outcomes included symptoms, HRQoL, fatigue, work productivity and activity impairment, assessed via validated PRO measures (respectively, EQ-5D-5L, PROMIS Fatigue 8a, WPAI:GH) and ad-hoc questionnaires, at different time points (Day 3, Week 1, Week 2, Week 4).

#### Acute symptoms

Using the CDC list of acute symptoms[12] as reference, the study assessed the presence of 12 symptoms at time of testing, Week 1, 2 and 4, across 3 categories: (1) systemic (including fever, chills, muscle or body aches, headaches, and fatigue) (2) respiratory (including shortness of breath or difficulty breathing, cough, sore throat, new/ recent loss of taste or smell); (3) gastrointestinal (GI) (including nausea or vomiting, and diarrhea). Symptom trajectories were reported as symptom point prevalence at each time point of the study. The pre-test screening questionnaire captured symptoms when participants scheduled their test, while an ad-hoc questionnaire including the same symptoms was administered at Weeks 1 and 2. At Week 4, an ad-hoc questionnaire was administered listing the 30 long-COVID symptoms including those from the CDC[13]. A total of 11 symptoms in this list matched the list of acute symptoms (all but ‘congestion or runny nose’) and were assessed and reported in these analyses.

#### Health-Related Quality of Life (HRQoL)

The study assessed HRQoL via the validated EQ-5D-5L questionnaire[14] that subjects were asked to complete at enrollment (Day 3), Week 2 and Week 4. Five dimensions of EQ-5D-5L at each time point were converted into the Utility Index (UI) using the US-based weights established by Pickard et al[15]. UI and visual analogue scale (VAS) scores were compared among cohorts and across assessment times. Lower scores for both EQ VAS and UI correspond to lower overall self-reported health-related quality of life.

#### Work Productivity and Activity Impairment

The Work Productivity and Activity Impairment General Health v2.0 (WPAI:GH) instrument was used to measure impairments in both paid and unpaid work [16,17]. Participants were asked to complete the survey at Week 1, Week 2 and Week 4. Higher scores correspond to greater activity impairment and work productivity loss. Only participants that reported being employed were included for work productivity analyses. WPAI results were compared among cohorts and across assessment times.

#### Fatigue

Fatigue was measured in the CDC-based symptoms questionnaire and also with the validated Patient Reported Outcomes Measurement Information System [PROMIS]Fatigue 8a, a short-form fixed instrument comprised of 8 of the 90-item PROMIS-Fatigue item bank[18,19]. Participants were asked to complete the survey at Week 1, Week 2 and Week 4. PROMIS uses T-scores, a type of standard score referenced to the US general population norms, which have a mean of 50 and standard deviation (SD) of 10. The raw summation score of 8 items was converted to standardized T-score based on the US population average [20]. Higher T-scores indicate worse fatigue[18].

### Statistical analysis

Means and standard deviations for continuous variables and frequency and percentages for categorical variables were used to summarize participant characteristics at baseline and outcomes at follow-up. T-tests and chi-square tests were used to test for continuous variables and categorical variables, respectively to measure between-group differences. Fisher’s exact tests were used for 2-by-2 tables and Fisher-Freeman-Halton tests for r-by-c tables when an expected cell frequency was less than 5 [21,22]. All *P* values were two-sided.

Mixed models for repeated measures (MMRM) were used to estimate the impact of vaccination on symptoms, HRQoL and WPAI over time[23]. Assessment time was fitted as a categorical covariate and a repeated effect (repeated by subject) with unstructured covariance matrix. Least squares mean (LS mean) and standard errors of PRO scores for each time point of assessment were calculated. Per guidelines, no adjustment was made for missing data when scoring the EQ-5D-5L Utility Index (UI) [15] and WPAI [17]. All available data were included in the analysis.

Cohen’s d, or a variation of it, was calculated to assess the difference in pre-COVID scores among patients vaccinated and unvaccinated, the magnitude of score change from baseline to each time point within each cohort, as well as the differences between cohorts (Bivalent vs. unvaccinated) [24,25]. Specifically, within-cohort effect size (ES) was calculated as mean change from baseline to follow-up, divided by the standard deviation of change scores from baseline to follow-up[4]. Between-cohort ES was calculated as the difference in mean score between cohorts, divided by the pooled standard deviation of scores or, alternatively, the difference in mean changes from baseline between cohorts, divided by the pooled standard deviation of change scores[4]. Values of 0.2, 0.5, and 0.8 standard deviation (SD) units represent “small,” “medium,” and “large” effect sizes, respectively[4,24].

On an exploratory basis, the two cohorts were matched on the clinical and demographic variables that differed, as an interim step between the analyses of the raw data and the MMRM. Matching results are presented in the Supplementary Material.

All analyses were conducted with SAS Version 9.4 (SAS Institute, Cary, NC). The study followed the Strengthening the Reporting of Observational Studies in Epidemiology (STROBE) reporting guideline[26].

## RESULTS

### Baseline characteristics

A total of 21,113 eligible candidates that tested positive at a CVS Health test site were outreached. Of these, 643 consented and met the inclusion and exclusion criteria for the analyses: 316 (49.1%) received bivalent BA.4/5 BNT162b2 and 327 (50.1%) were unvaccinated (Figure 1). Compared with individuals in the CVS Health analytic dataset who did not participate in our study, the study sample was slightly older, over-represented by women and Caucasians, with slightly more comorbidities and lower SVI. The study sample reported a similar number of acute symptoms during the infection, although a higher proportion of those in the study reported cough, new loss of taste/smell, nausea, or vomiting, and a lower proportion reported diarrhea and muscle or body aches (Supplemental Table 1).

Baseline sociodemographic characteristics of the participants are shown in Table 1. Overall, the mean (SD) age was 46.5 (15.9), 70.3% were female, 58.2% Caucasian, and 40.4% were from the Southern US. The sample was characterized by moderate social vulnerability (mean SVI: 0.446). Almost half (44.2%) reported a previous COVID-19 infection, 25.7% reported at least 1 comorbidity, and 24.3% reported being prescribed a COVID-19 antiviral for the current infection. Compared with unvaccinated, bivalent BA.4/5 BNT162b2 participants were comparable with respect to sex. However, they tended to be older, Caucasian, reside in the Southern US, have lower SVI, have more comorbidities and utilize COVID-19 antivirals. In the vaccinated group, mean (SD) time between bivalent vaccination and infection was 165 (SD: 45) days. At Week 4, 72 (22.8%) and 100 (30.6%) participants did not respond to the assigned surveys, respectively, in the bivalent and unvaccinated cohorts. After matching for age, race/ethnicity, SVI category, region, >=1 comorbidity, and antiviral use, the patient characteristics between the two groups were balanced (Supplemental Table 2).

**Table 1:** Baseline characteristics stratified by vaccination status.

|  | All | BNT162b2 | Unvaccinated | *P*<sup>a</sup> |
| --- | --- | --- | --- | --- |
| Total, n (%) | 643 (100.0) | 316 (49.1) | 327 (50.9) |  |
| Age, years |  |  |  |  |
| Mean, SD | 46.5 (15.9) | 50.8 (16.0) | 42.3 (14.7) | 0.000 |
| Age group, n (%) |  |  |  | 0.000 |
| 18-29 | 109 (17.0) | 33 (10.4) | 76 (23.2) |  |
| 30-49 | 257 (40.0) | 111 (35.1) | 146 (44.6) |  |
| 50-64 | 167 (26.0) | 93 (29.4) | 74 (22.6) |  |
| 65-74 | 89 (13.8) | 62 (19.6) | 27 (8.3) |  |
| ≥75 | 21 (3.2) | 17 (5.4) | 4 (1.2) |  |
| Gender, n (%) |  |  |  | 0.185 |
| Female | 452 (70.3) | 212 (67.1) | 240 (73.4) |  |
| Male | 185 (28.8) | 101 (32.0) | 84 (25.7) |  |
| Unknown | 6 (0.9) | 3 (1.0) | 3 (0.9) |  |
| Race / Ethnicity, n (%) |  |  |  | 0.000 |
| White or Caucasian (not Hispanic or Latino) | 374 (58.2) | 204 (64.6) | 170 (52.0) |  |
| Black or African American | 57 (8.9) | 19 (6.0) | 38 (11.6) |  |
| Hispanic | 99 (15.4) | 33 (10.4) | 66 (20.2) |  |
| Asian | 63 (9.8) | 36 (11.4) | 27 (8.3) |  |
| Patient Refused | 22 (3.4) | 14 (4.4) | 8 (2.5) |  |
| Other | 28 (4.3) | 10 (3.2) | 18 (5.5) |  |
| US Geographic Region, n (%) |  |  |  | 0.013 |
| Northeast | 88 (13.7) | 51 (16.1) | 37 (11.3) |  |
| South | 260 (40.4) | 108 (34.2) | 152 (46.5) |  |
| Midwest | 141 (21.9) | 76 (24.1) | 65 (19.9) |  |
| West | 154 (24.0) | 81 (25.6) | 73 (22.3) |  |
| CMS Geographic Region (n, %) |  |  |  | 0.001 |
| Region 1: ME, NH, VT, MA, CT, RI | 43 (6.7) | 27 (8.5) | 16 (4.9) |  |
| Region 2: NY, NJ, PR, VI | 21 (3.3) | 11 (3.5) | 10 (3.1) |  |
| Region 3: PA, DE, MD, DC, WV, VA | 52 (8.1) | 31 (9.8) | 21 (6.4) |  |
| Region 4: KY, TN, NC, SC, GA, MS, AL, FL | 146 (22.7) | 63 (19.9) | 83 (25.4) |  |
| Region 5: MN, WI, IL, MI, IN, OH | 132 (20.5) | 70 (22.2) | 62 (19.0) |  |
| Region 6: NM, OK, AR, TX, LA | 88 (13.7) | 28 (8.9) | 60 (18.3) |  |
| Region 7: NE, IA, KS, MO | 9 (1.4) | 6 (1.9) | 3 (0.9) |  |
| Region 8: MT, ND, SD, WY, UT, CO | 5 (0.8) | 4 (1.3) | 1 (0.3) |  |
| Region 9: CA, NV, AZ, GU | 142 (22.1) | 71 (22.5) | 71 (21.7) |  |
| Region 10: AK, WA, OR, ID | 5 (0.8) | 5 (1.6) | 0 (0.0) |  |
| Social vulnerability index, Mean (SD)<sup>b</sup> | 0.45 (0.2) | 0.39 (0.2) | 0.50 (0.2) | 0.000 |
| Previously Tested Positive, n (%) |  |  |  | 0.051 |
| No | 338 (52.6) | 177 (56.0) | 161 (49.2) |  |
| Yes | 268 (41.7) | 119 (37.7) | 149 (45.6) |  |
| Missing | 37 (5.8) | 20 (6.3) | 17 (5.3) |  |
| Number of comorbidities, Mean (SD) | 0.40 (0.8) | 0.47 (0.8) | 0.34 (0.8) | 0.043 |
| At least 1 comorbidity, n (%) | 165 (25.7) | 96 (30.4) | 69 (21.1) | 0.007 |
| Self-Reported Comorbidity, n (%) |  |  |  |  |
| Asthma or Chronic Lung Disease | 33 (5.1) | 20 (6.3) | 13 (4.0) | 0.176 |
| Cirrhosis of the liver | 2 (0.3) | 0 (0.0) | 2 (0.6) | 0.499 |
| Immunocompromised Conditions or Weakened Immune System<sup>c</sup> | 2 (0.3) | 1 (0.3) | 1 (0.3) | 1.000 |
| Diabetes | 39 (6.1) | 23 (7.3) | 16 (4.9) | 0.205 |
| Heart Conditions or Hypertension | 104 (16.2) | 60 (19.0) | 44 (13.5) | 0.057 |
| Overweight or obesity | 80 (12.4) | 44 (13.9) | 36 (11.0) | 0.263 |
| Paxlovid prescription, n (%) |  |  |  | 0.000 |
| No | 493 (76.7) | 222 (70.3) | 271 (82.9) |  |
| Yes | 148 (23.0) | 92 (29.1) | 56 (17.1) |  |
| Missing | 2 (0.3) | 2 (0.6) | 0 (0.0) |  |

CMS: Centers for Medicare and Medicaid Services; SD: Standard Deviation
<sup>a</sup> *P* value refers to the comparison between BNT162b2 and Unvaccinated.
<sup>b</sup> SVI ranges from 0 to 1. A community with higher value is more socially vulnerable.
<sup>c</sup> Immunocompromised conditions includes compromised immune system (such as from immune-compromising drugs, solid organ or blood stem cell transplant, HIV, or other conditions), conditions that result in a weakened immune system, including kidney failure or end stage renal disease

### Acute symptoms

At the time of testing, study participants reported a mean of 5.3 symptoms, with the most frequent being respiratory and systemic symptoms (Table 2). BA.4/5 BNT162b2 participants reported fewer overall acute COVID-19 symptoms than unvaccinated: a mean of 5.0 vs. 5.7 (p<0.001). The proportions of systemic symptoms were lower in the BA.4/5 BNT162b2 cohort, driven by lower frequency of fever, chills, muscle or body aches, and headaches. All the other symptoms were directionally less frequent in the BA.4/5 BNT162b2 cohort (Table 2). These results were generally consistent based on the (repeated measures) model and after matching (Table 2, Supplemental Table 3). Supplemental Figure 2 shows the prevalence of individual symptoms by exposure.

**Table 2:**
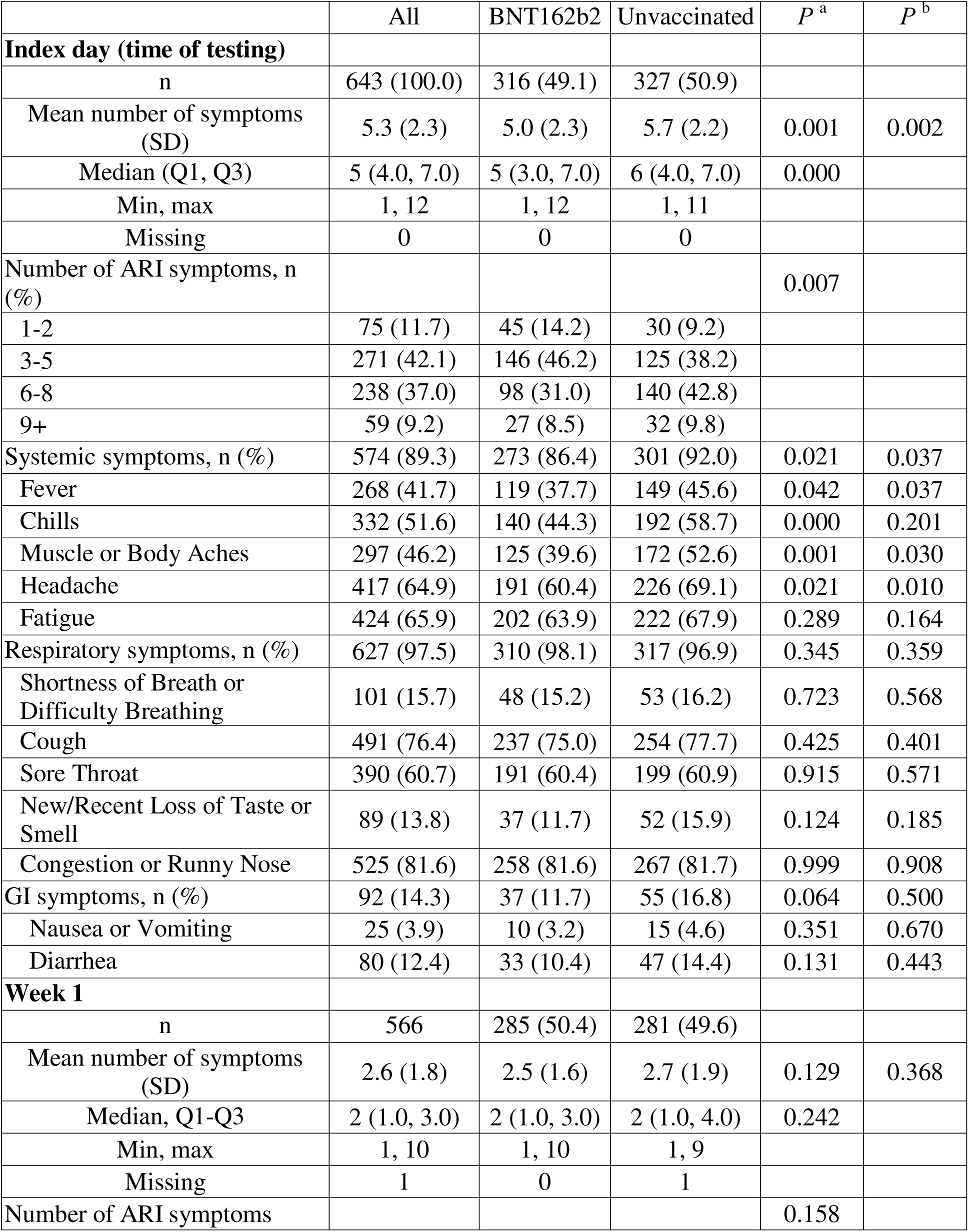
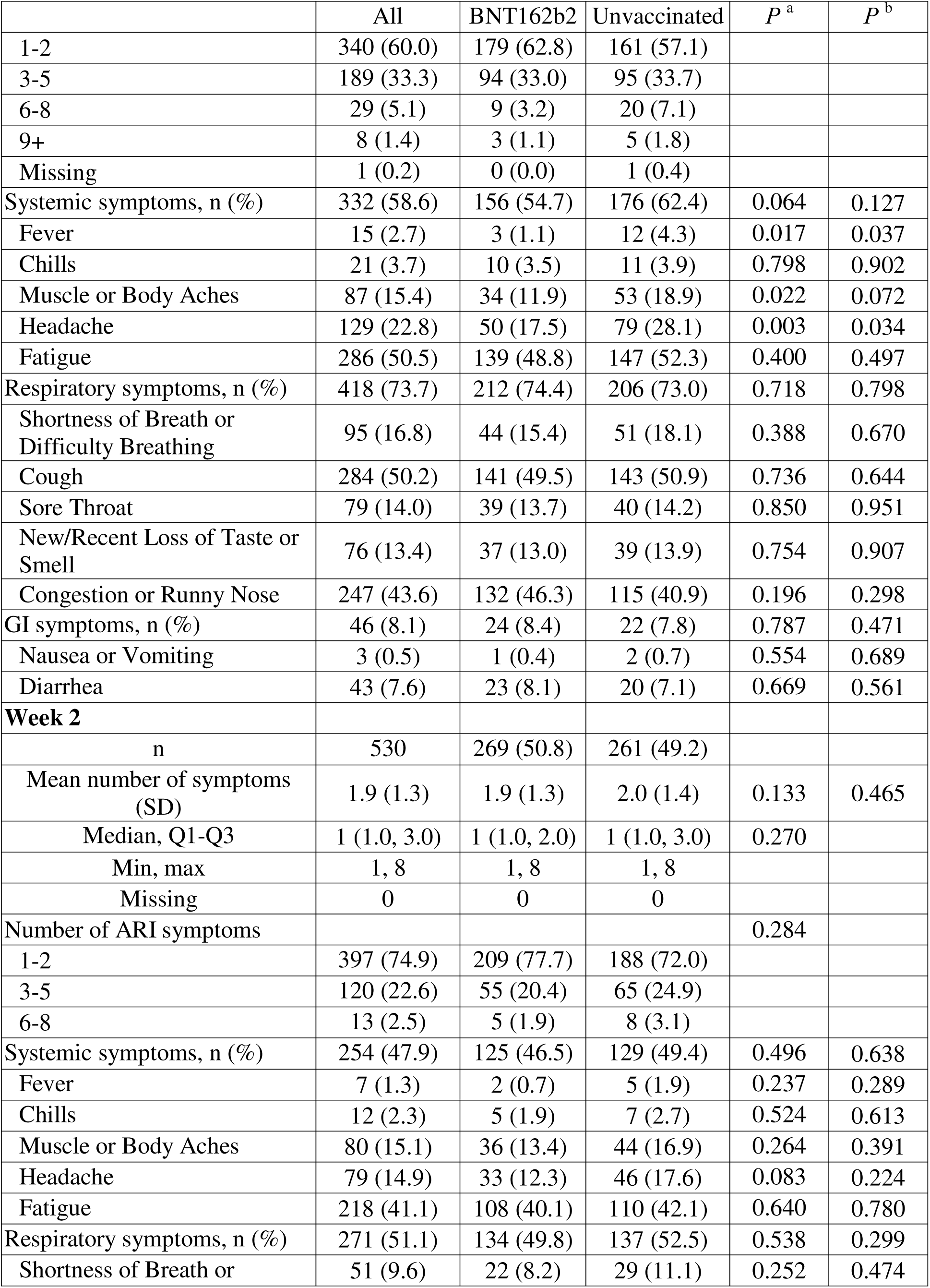
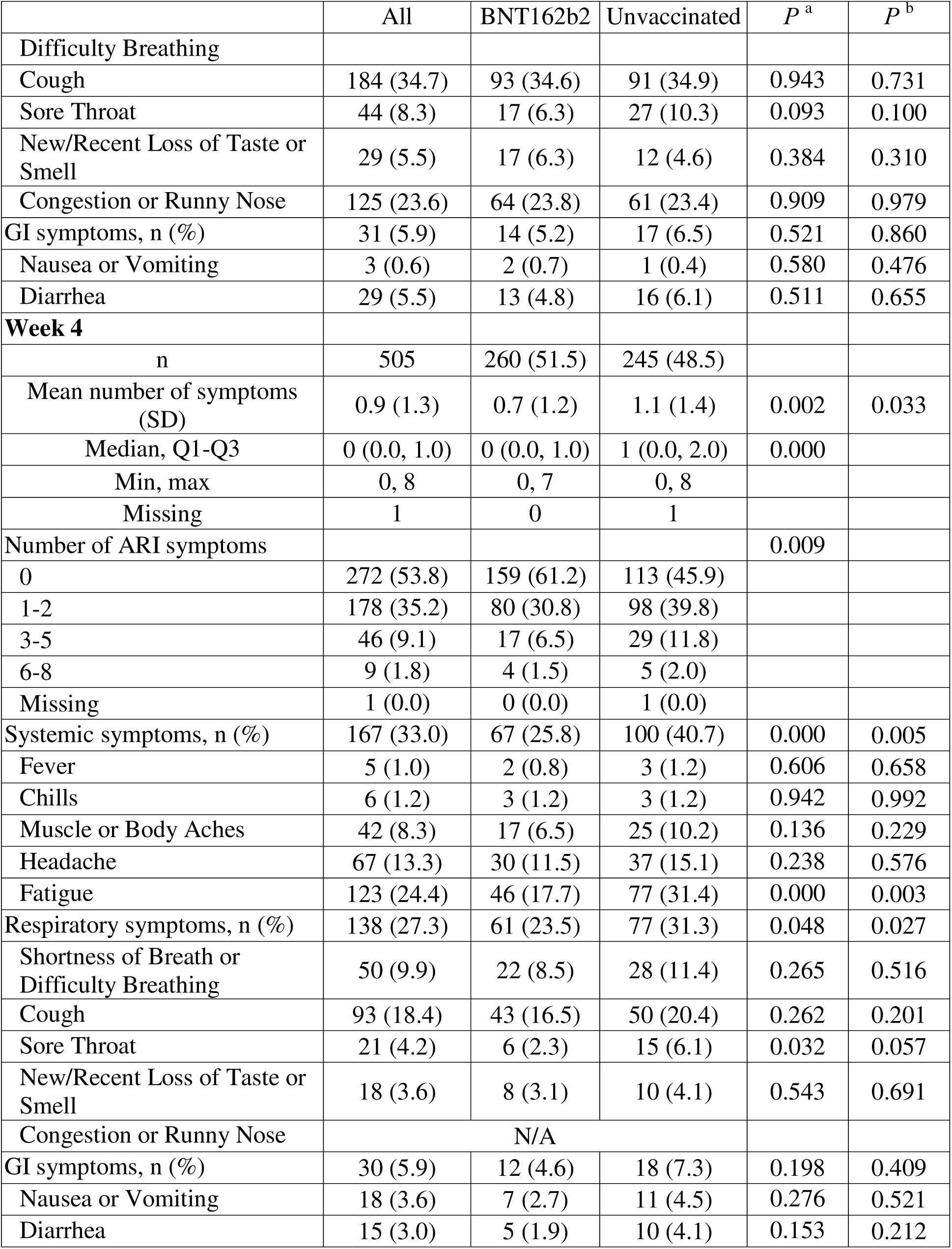
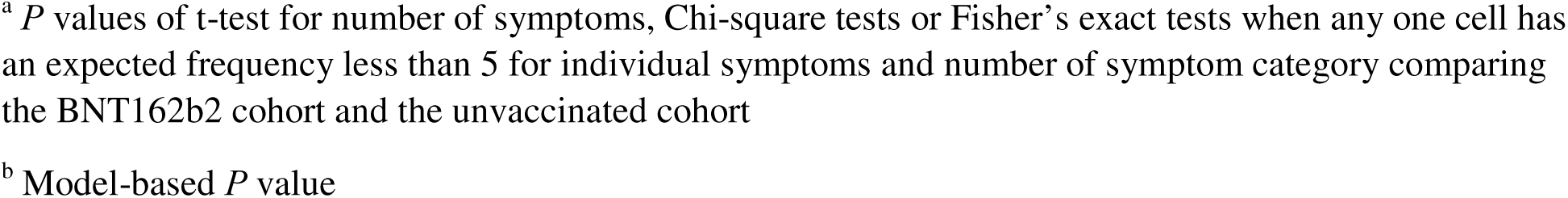
Trajectory of acute COVID-19 symptoms at time of testing, Week 1, Week 2, and Week 4.

At Week 1, the number of symptoms halved, dropping to a mean of 2.6 (Table 2). The BA.4/5 BNT162b2 cohort had a numerically lower, although not statistically significant, mean number of symptoms (2.5 vs. 2.7, p=0.129) driven by fewer systemic symptoms (fever, muscle or body ache, headache). The other symptoms were directionally less frequent in the BA.4/5 BNT162b2 cohort, except for congestion, runny nose, and diarrhea. These results were generally consistent after matching (Supplemental Table 3); the model-based results showed that vaccinated participants had less fever and headache (Table 2).

At Week 2, the mean number of symptoms dropped to 1.9 (Table 2). Except for loss of taste or smell, congestion or runny nose, and nausea or vomiting, symptoms were directionally less frequent in the BA.4/5 BNT162b2 cohort. These results were consistent based on the model (Table 2) and matching (Supplemental Table 3).

At Week 4, the mean number of acute symptoms dropped to 0.9 (Table 2). BA.4/5 BNT162b2 participants reported fewer symptoms than unvaccinated (mean: 0.7 vs. 1.1, p=0.002), driven by less systemic symptoms and respiratory symptoms. Fatigue and sore throat were less prevalent in the BA.4/5 BNT162b2 cohort. After matching, the point prevalence of muscle or body aches was also lower in the bivalent cohort (Supplemental Table 3). All the other symptoms were numerically less frequent in the BA.4/5 BNT162b2 cohort (Table 2).

When stratifying participants by ordinal categories of self-reported symptoms (0, 1-2, 3-5, 6+) and vaccination status, the BA.4/5 BNT162b2 cohort was characterized by lower proportions of participants with high symptom burden compared with unvaccinated across all time points and to a greater extent at the time of testing and Week 4 (Figure 2).

**Figure 2.**
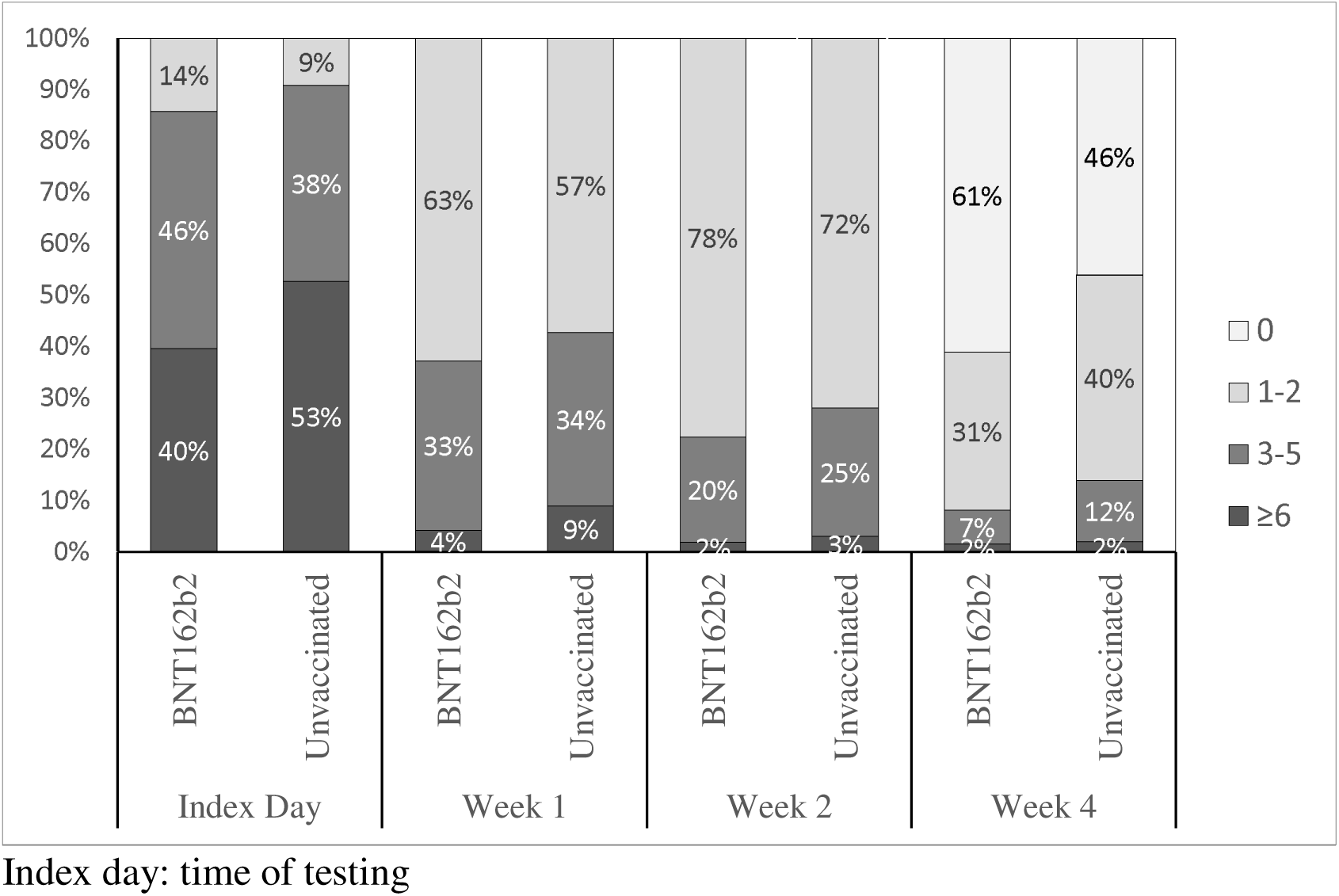
Distribution of study participants across ordinal categories of number of symptoms and vaccination status, across time points.

### Health-Related Quality of life

The mean pre-COVID-19 UIs did not differ between the bivalent BA.4/5 BNT162b2 and unvaccinated cohorts, respectively 0.930 and 0.928 (P=0.804) (Table 3). COVID-19 had a detrimental effect on the HRQoL of participants, especially during Day 3. In both the BA.4/5 BNT162b2 and the unvaccinated cohorts, UIs were lower at Day 3, Week 2 and 4 relative to pre-COVID-19. While improvement was observed over time, neither the observed nor the model-based UI scores returned to pre-COVID levels at Week 4. The observed and model-based UIs were numerically higher in the BA.4/5 BNT162b2 cohort across all time points but were not significantly different from those in the unvaccinated cohort (Table 3, Table 4). Mean pre-COVID-19 EQ-VAS scores were similar for the BA.4/5 BNT162b2 and unvaccinated cohorts, respectively 85.8 and 86.3 (P=0.581) (Table 3). The observed and model-based UIs were numerically lower in the BA.4/5 BNT162b2 cohort across all time points and were not significantly different from those in the unvaccinated cohort. These results were generally similar post-matching (Supplemental Table 5).

**Table 3:**
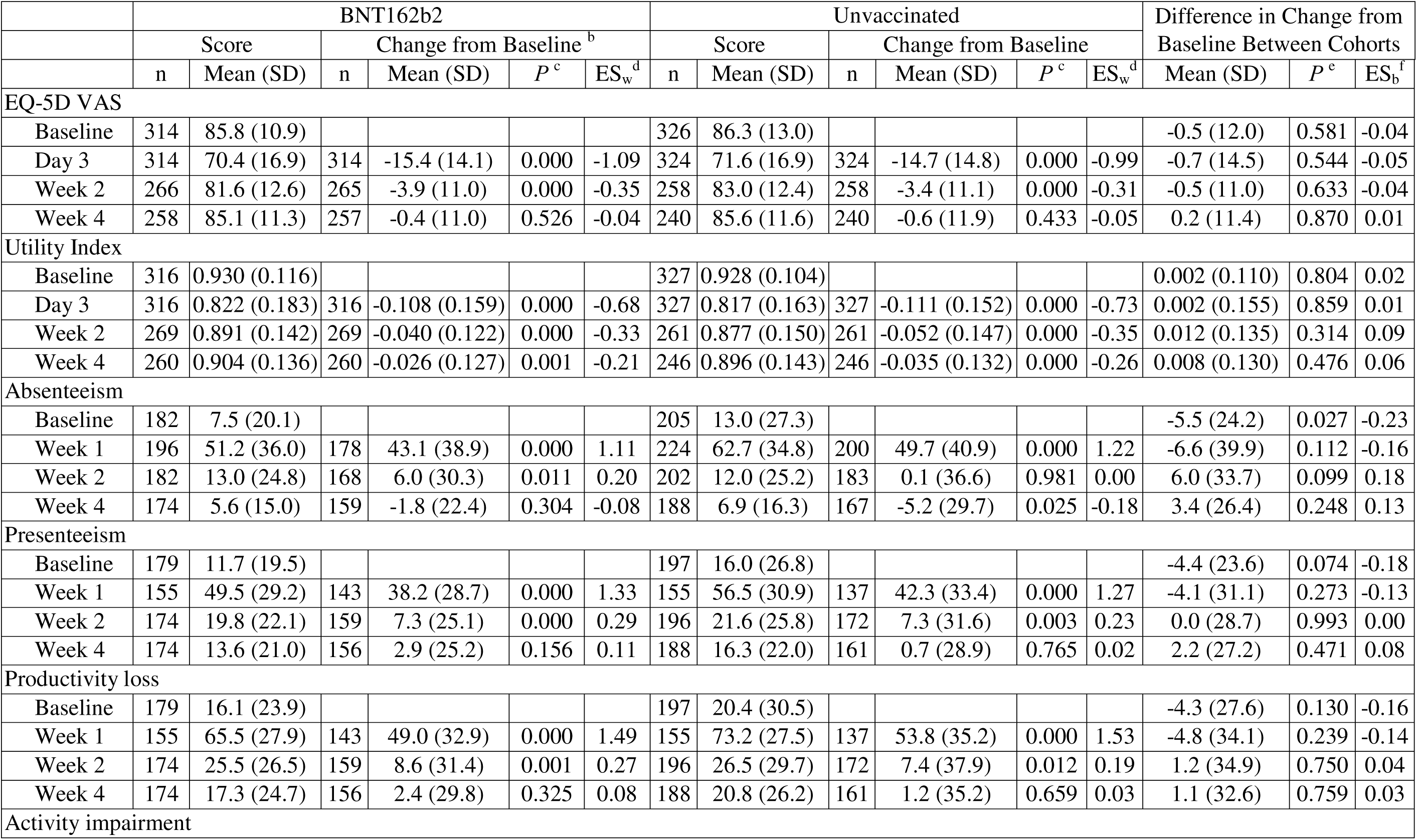
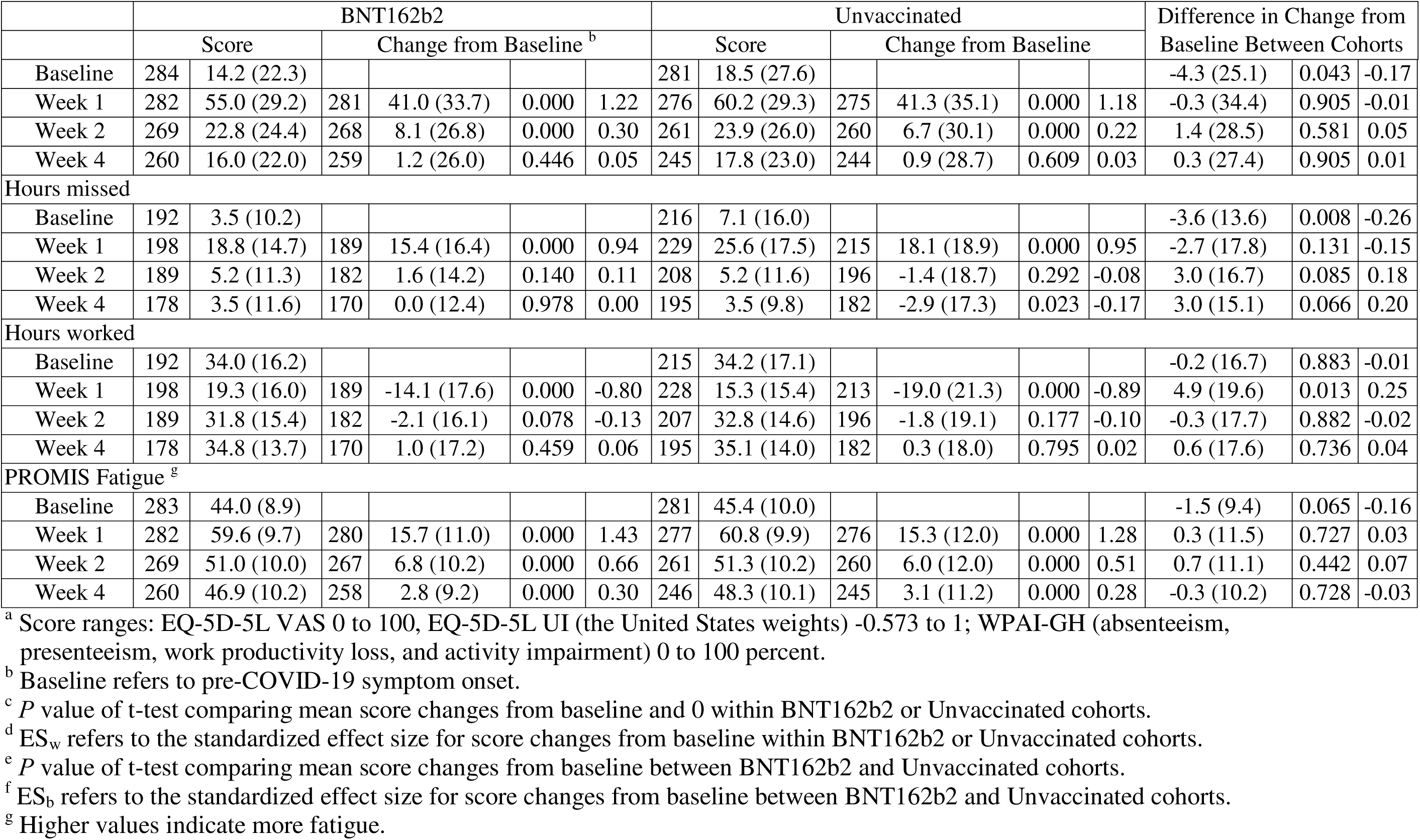
Summary of Observed EQ-5D-5L, PROMIS Fatigue and WPAI-GH Scores ^a^ and Their Changes from Baseline by Assessment Time and Vaccination Status.

|  | BNT162b2 |  |  |  |  |  | Unvaccinated |  |  |  |  |  | Difference in Change from Baseline Between Cohorts |  |  |
| --- | --- | --- | --- | --- | --- | --- | --- | --- | --- | --- | --- | --- | --- | --- | --- |
|  | Score |  | Change from Baseline<sup>b</sup> |  |  |  | Score |  | Change from Baseline |  |  |  |  |  |  |
|  | n | Mean (SD) | n | Mean (SD) | *P*<sup>c</sup> | ES<sub>w</sub><sup>d</sup> | n | Mean (SD) | n | Mean (SD) | *P*<sup>c</sup> | ES<sub>w</sub><sup>d</sup> | Mean (SD) | *P*<sup>e</sup> | ES<sub>b</sub><sup>f</sup> |
| EQ-5D VAS |  |  |  |  |  |  |  |  |  |  |  |  |  |  |  |
| Baseline | 314 | 85.8 (10.9) |  |  |  |  | 326 | 86.3 (13.0) |  |  |  |  | -0.5 (12.0) | 0.581 | -0.04 |
| Day 3 | 314 | 70.4 (16.9) | 314 | -15.4 (14.1) | 0.000 | -1.09 | 324 | 71.6 (16.9) | 324 | -14.7 (14.8) | 0.000 | -0.99 | -0.7 (14.5) | 0.544 | -0.05 |
| Week 2 | 266 | 81.6 (12.6) | 265 | -3.9 (11.0) | 0.000 | -0.35 | 258 | 83.0 (12.4) | 258 | -3.4 (11.1) | 0.000 | -0.31 | -0.5 (11.0) | 0.633 | -0.04 |
| Week 4 | 258 | 85.1 (11.3) | 257 | -0.4 (11.0) | 0.526 | -0.04 | 240 | 85.6 (11.6) | 240 | -0.6 (11.9) | 0.433 | -0.05 | 0.2 (11.4) | 0.870 | 0.01 |
| Utility Index |  |  |  |  |  |  |  |  |  |  |  |  |  |  |  |
| Baseline | 316 | 0.930 (0.116) |  |  |  |  | 327 | 0.928 (0.104) |  |  |  |  | 0.002 (0.110) | 0.804 | 0.02 |
| Day 3 | 316 | 0.822 (0.183) | 316 | -0.108 (0.159) | 0.000 | -0.68 | 327 | 0.817 (0.163) | 327 | -0.111 (0.152) | 0.000 | -0.73 | 0.002 (0.155) | 0.859 | 0.01 |
| Week 2 | 269 | 0.891 (0.142) | 269 | -0.040 (0.122) | 0.000 | -0.33 | 261 | 0.877 (0.150) | 261 | -0.052 (0.147) | 0.000 | -0.35 | 0.012 (0.135) | 0.314 | 0.09 |
| Week 4 | 260 | 0.904 (0.136) | 260 | -0.026 (0.127) | 0.001 | -0.21 | 246 | 0.896 (0.143) | 246 | -0.035 (0.132) | 0.000 | -0.26 | 0.008 (0.130) | 0.476 | 0.06 |
| Absenteeism |  |  |  |  |  |  |  |  |  |  |  |  |  |  |  |
| Baseline | 182 | 7.5 (20.1) |  |  |  |  | 205 | 13.0 (27.3) |  |  |  |  | -5.5 (24.2) | 0.027 | -0.23 |
| Week 1 | 196 | 51.2 (36.0) | 178 | 43.1 (38.9) | 0.000 | 1.11 | 224 | 62.7 (34.8) | 200 | 49.7 (40.9) | 0.000 | 1.22 | -6.6 (39.9) | 0.112 | -0.16 |
| Week 2 | 182 | 13.0 (24.8) | 168 | 6.0 (30.3) | 0.011 | 0.20 | 202 | 12.0 (25.2) | 183 | 0.1 (36.6) | 0.981 | 0.00 | 6.0 (33.7) | 0.099 | 0.18 |
| Week 4 | 174 | 5.6 (15.0) | 159 | -1.8 (22.4) | 0.304 | -0.08 | 188 | 6.9 (16.3) | 167 | -5.2 (29.7) | 0.025 | -0.18 | 3.4 (26.4) | 0.248 | 0.13 |
| Presenteeism |  |  |  |  |  |  |  |  |  |  |  |  |  |  |  |
| Baseline | 179 | 11.7 (19.5) |  |  |  |  | 197 | 16.0 (26.8) |  |  |  |  | -4.4 (23.6) | 0.074 | -0.18 |
| Week 1 | 155 | 49.5 (29.2) | 143 | 38.2 (28.7) | 0.000 | 1.33 | 155 | 56.5 (30.9) | 137 | 42.3 (33.4) | 0.000 | 1.27 | -4.1 (31.1) | 0.273 | -0.13 |
| Week 2 | 174 | 19.8 (22.1) | 159 | 7.3 (25.1) | 0.000 | 0.29 | 196 | 21.6 (25.8) | 172 | 7.3 (31.6) | 0.003 | 0.23 | 0.0 (28.7) | 0.993 | 0.00 |
| Week 4 | 174 | 13.6 (21.0) | 156 | 2.9 (25.2) | 0.156 | 0.11 | 188 | 16.3 (22.0) | 161 | 0.7 (28.9) | 0.765 | 0.02 | 2.2 (27.2) | 0.471 | 0.08 |
| Productivity loss |  |  |  |  |  |  |  |  |  |  |  |  |  |  |  |
| Baseline | 179 | 16.1 (23.9) |  |  |  |  | 197 | 20.4 (30.5) |  |  |  |  | -4.3 (27.6) | 0.130 | -0.16 |
| Week 1 | 155 | 65.5 (27.9) | 143 | 49.0 (32.9) | 0.000 | 1.49 | 155 | 73.2 (27.5) | 137 | 53.8 (35.2) | 0.000 | 1.53 | -4.8 (34.1) | 0.239 | -0.14 |
| Week 2 | 174 | 25.5 (26.5) | 159 | 8.6 (31.4) | 0.001 | 0.27 | 196 | 26.5 (29.7) | 172 | 7.4 (37.9) | 0.012 | 0.19 | 1.2 (34.9) | 0.750 | 0.04 |
| Week 4 | 174 | 17.3 (24.7) | 156 | 2.4 (29.8) | 0.325 | 0.08 | 188 | 20.8 (26.2) | 161 | 1.2 (35.2) | 0.659 | 0.03 | 1.1 (32.6) | 0.759 | 0.03 |
| Activity impairment |  |  |  |  |  |  |  |  |  |  |  |  |  |  |  |

|  | BNT162b2 |  |  |  |  |  | Unvaccinated |  |  |  |  |  | Difference in Change from<br>Baseline Between Cohorts |  |  |
| --- | --- | --- | --- | --- | --- | --- | --- | --- | --- | --- | --- | --- | --- | --- | --- |
|  | Score |  | Change from Baseline <sup>b</sup> |  |  |  | Score |  | Change from Baseline |  |  |  |  |  |  |
| Baseline | 284 | 14.2 (22.3) |  |  |  |  | 281 | 18.5 (27.6) |  |  |  |  | -4.3 (25.1) | 0.043 | -0.17 |
| Week 1 | 282 | 55.0 (29.2) | 281 | 41.0 (33.7) | 0.000 | 1.22 | 276 | 60.2 (29.3) | 275 | 41.3 (35.1) | 0.000 | 1.18 | -0.3 (34.4) | 0.905 | -0.01 |
| Week 2 | 269 | 22.8 (24.4) | 268 | 8.1 (26.8) | 0.000 | 0.30 | 261 | 23.9 (26.0) | 260 | 6.7 (30.1) | 0.000 | 0.22 | 1.4 (28.5) | 0.581 | 0.05 |
| Week 4 | 260 | 16.0 (22.0) | 259 | 1.2 (26.0) | 0.446 | 0.05 | 245 | 17.8 (23.0) | 244 | 0.9 (28.7) | 0.609 | 0.03 | 0.3 (27.4) | 0.905 | 0.01 |
| Hours missed |  |  |  |  |  |  |  |  |  |  |  |  |  |  |  |
| Baseline | 192 | 3.5 (10.2) |  |  |  |  | 216 | 7.1 (16.0) |  |  |  |  | -3.6 (13.6) | 0.008 | -0.26 |
| Week 1 | 198 | 18.8 (14.7) | 189 | 15.4 (16.4) | 0.000 | 0.94 | 229 | 25.6 (17.5) | 215 | 18.1 (18.9) | 0.000 | 0.95 | -2.7 (17.8) | 0.131 | -0.15 |
| Week 2 | 189 | 5.2 (11.3) | 182 | 1.6 (14.2) | 0.140 | 0.11 | 208 | 5.2 (11.6) | 196 | -1.4 (18.7) | 0.292 | -0.08 | 3.0 (16.7) | 0.085 | 0.18 |
| Week 4 | 178 | 3.5 (11.6) | 170 | 0.0 (12.4) | 0.978 | 0.00 | 195 | 3.5 (9.8) | 182 | -2.9 (17.3) | 0.023 | -0.17 | 3.0 (15.1) | 0.066 | 0.20 |
| Hours worked |  |  |  |  |  |  |  |  |  |  |  |  |  |  |  |
| Baseline | 192 | 34.0 (16.2) |  |  |  |  | 215 | 34.2 (17.1) |  |  |  |  | -0.2 (16.7) | 0.883 | -0.01 |
| Week 1 | 198 | 19.3 (16.0) | 189 | -14.1 (17.6) | 0.000 | -0.80 | 228 | 15.3 (15.4) | 213 | -19.0 (21.3) | 0.000 | -0.89 | 4.9 (19.6) | 0.013 | 0.25 |
| Week 2 | 189 | 31.8 (15.4) | 182 | -2.1 (16.1) | 0.078 | -0.13 | 207 | 32.8 (14.6) | 196 | -1.8 (19.1) | 0.177 | -0.10 | -0.3 (17.7) | 0.882 | -0.02 |
| Week 4 | 178 | 34.8 (13.7) | 170 | 1.0 (17.2) | 0.459 | 0.06 | 195 | 35.1 (14.0) | 182 | 0.3 (18.0) | 0.795 | 0.02 | 0.6 (17.6) | 0.736 | 0.04 |
| PROMIS Fatigue <sup>g</sup> |  |  |  |  |  |  |  |  |  |  |  |  |  |  |  |
| Baseline | 283 | 44.0 (8.9) |  |  |  |  | 281 | 45.4 (10.0) |  |  |  |  | -1.5 (9.4) | 0.065 | -0.16 |
| Week 1 | 282 | 59.6 (9.7) | 280 | 15.7 (11.0) | 0.000 | 1.43 | 277 | 60.8 (9.9) | 276 | 15.3 (12.0) | 0.000 | 1.28 | 0.3 (11.5) | 0.727 | 0.03 |
| Week 2 | 269 | 51.0 (10.0) | 267 | 6.8 (10.2) | 0.000 | 0.66 | 261 | 51.3 (10.2) | 260 | 6.0 (12.0) | 0.000 | 0.51 | 0.7 (11.1) | 0.442 | 0.07 |
| Week 4 | 260 | 46.9 (10.2) | 258 | 2.8 (9.2) | 0.000 | 0.30 | 246 | 48.3 (10.1) | 245 | 3.1 (11.2) | 0.000 | 0.28 | -0.3 (10.2) | 0.728 | -0.03 |

<sup>a</sup> Score ranges: EQ-5D-5L VAS 0 to 100, EQ-5D-5L UI (the United States weights) -0.573 to 1; WPAI-GH (absenteeism, presenteeism, work productivity loss, and activity impairment) 0 to 100 percent.
<sup>b</sup> Baseline refers to pre-COVID-19 symptom onset.
<sup>c</sup> *P* value of t-test comparing mean score changes from baseline and 0 within BNT162b2 or Unvaccinated cohorts.
<sup>d</sup> *ES<sub>w</sub>* refers to the standardized effect size for score changes from baseline within BNT162b2 or Unvaccinated cohorts.
<sup>e</sup> *P* value of t-test comparing mean score changes from baseline between BNT162b2 and Unvaccinated cohorts.
<sup>f</sup> *ES<sub>b</sub>* refers to the standardized effect size for score changes from baseline between BNT162b2 and Unvaccinated cohorts.
<sup>g</sup> Higher values indicate more fatigue.

**Table 4.**
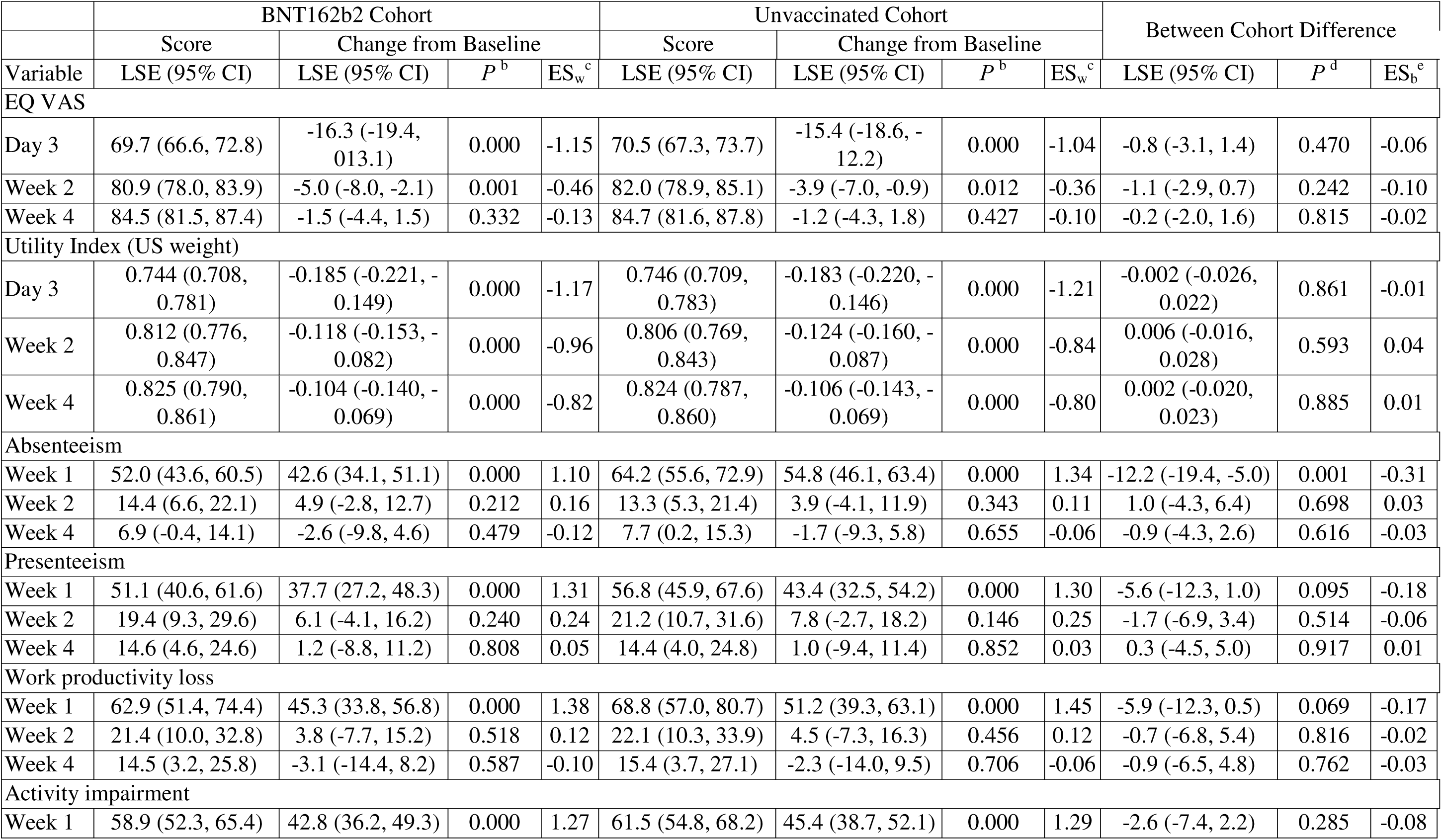
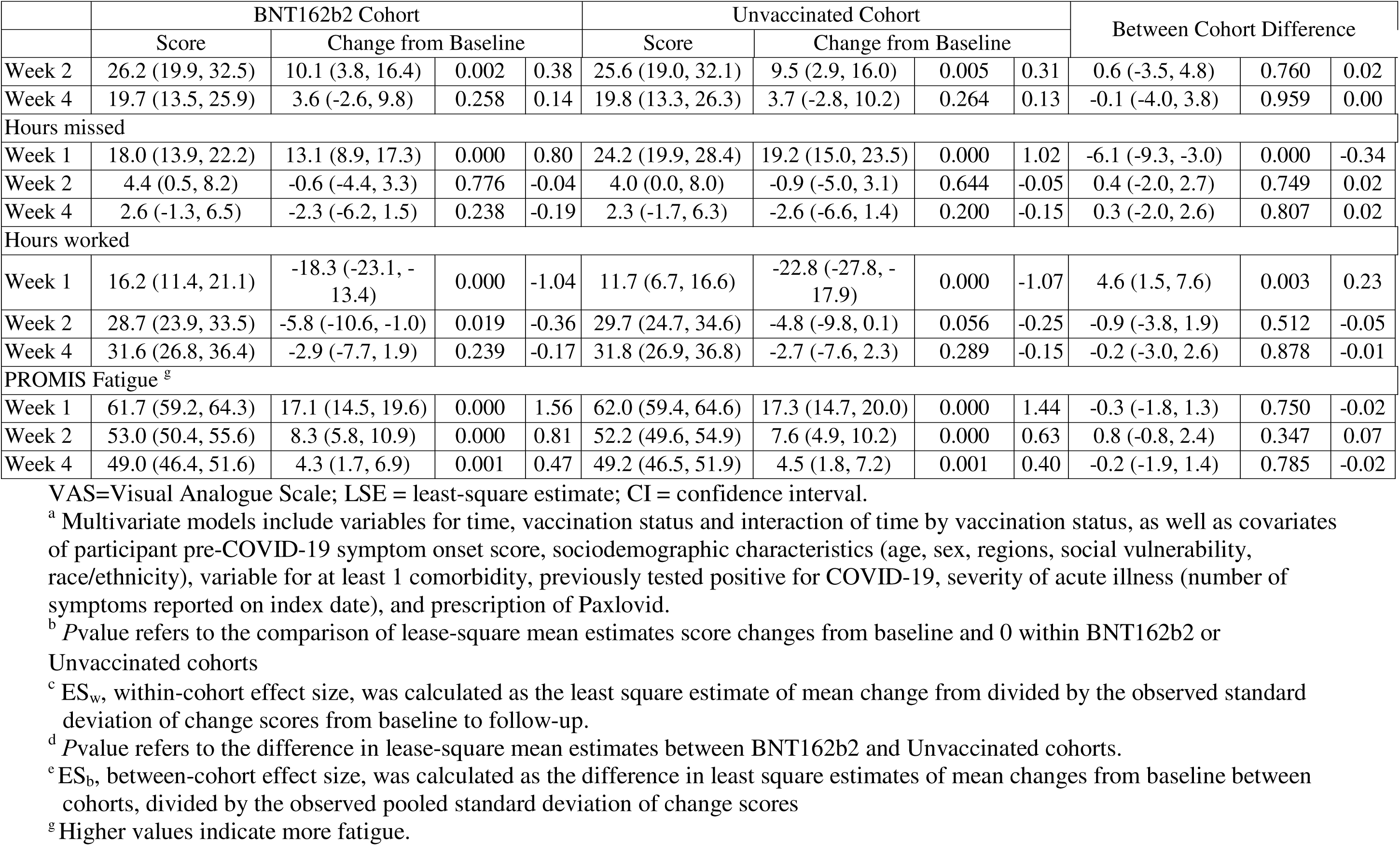
Least-Square Mean Estimates of EQ-5D-5L, PROMIS Fatigue and WPAI-GH Scores and Their Changes from Baseline by Assessment Time and Vaccination Status^a^.

### PROMIS Fatigue

The mean pre-COVID-19 baseline fatigue scores did not differ between the BA.4/5 BNT162b2 and unvaccinated cohorts, respectively 44.0 and 45.4 (P=0.065) (Table 3). COVID-19 infection had a detrimental effect on fatigue of participants, especially at Week 1 (T-score=60.2, indicating moderate fatigue). In both the BA.4/5 BNT162b2 and the unvaccinated cohorts, the observed and model-based scores were lower at all time points compared to pre-COVID-19. While improvement was observed over time, neither the observed nor the model-based scores returned to pre-COVID levels at Week 4 (Table 3, Table 4). No significant differences were observed between the two cohorts. Results were generally similar post-matching (Supplemental Table 5).

### Work Productivity and Activity Impairment

Approximately 74.1% of participants reported being employed at the time of testing (196 (62%) in the BA.4/5 BNT162b2 cohort and 224 (69%) unvaccinated), hence eligible to complete the absenteeism, presenteeism and work-productivity loss questions of WPAI:GH.

The mean pre-COVID-19 presenteeism and work productivity scores did not differ between the bivalent BA.4/5 BNT162b2 and unvaccinated cohorts, while absenteeism and activity impairment scores were slightly lower for the bivalent BA.4/5 BNT162b2 cohort (Table 3). COVID-19 infection negatively affected all WPAI dimensions, especially during Week 1 (Table 3, Table 4).

The absenteeism score was lower among vaccinated than unvaccinated at Week 1 (52.0% versus 64.2%), with a moderate ES of 0.31. In both cohorts, absenteeism returned to levels comparable to pre-COVID at Week 2 (Table 4). The presenteeism and work productivity scores returned to levels comparable to pre-COVID at Week 2 in both cohorts. The observed and model-based scores for these two WPAI domains were numerically lower among vaccinated, although not significantly different (Table 4). The bivalent cohort reported significantly fewer work hours missed during Week 1 versus the unvaccinated cohort (LSE mean: 18.0 versus 24.2); consistently, they reported more work hours worked (LSE mean: 16.2 versus 11.7) (Table 4).

The nonwork-related activity impairment scores returned to levels comparable to pre-COVID at Week 4 in both cohorts; scores were numerically lower among vaccinated, although not significantly different. The WPAI results were generally similar after matching (Supplemental Table 5). The model parameter estimates are presented in Supplemental Table 4.

## DISCUSSION

This nationwide study estimated the impact of the Pfizer-BioNTech BNT162b2 BA.4/5 bivalent vaccine on acute COVID-19 symptoms, Health-Related Quality of Life, Fatigue and Work Productivity and Activity Impairment among symptomatic adults testing positive for SARS-CoV-2 at a large national US pharmacy chain during circulation of the XBB 1.5 Omicron sub-lineage. Overall, the study found that COVID-19 infection had a detrimental effect on all patient outcomes, especially during Week 1, resulting in prolonged limitation of activities and of work productivity. However, the bivalent BA.4/5 BNT162b2 cohort was associated with significantly fewer and less persistent acute systemic and respiratory symptoms at the time of testing and Week 4. Across all time points, the BA.4/5 BNT162b2 cohort was characterized by lower proportions of participants with high symptom burden compared to unvaccinated/not up-to-date. These findings suggest that BA.4/5 BNT162b2 could alleviate the severity of infection, as measured by symptoms, and support patient recovery after SARS-CoV-2 infection.

Our findings on acute symptoms are concordant with the results of our prior study[4] that employed a similar design to assess the impact of monovalent BNT162b2 on the same acute symptoms and PROs, recruiting subjects a year earlier, during the first quarter of 2022. The study populations shared key similarities in characteristics, frequency and prevalence of acute symptoms at time of testing, and pre-COVID EQ-5D-5L and WPAI scores. In both studies, BNT162b2 COVID-19 vaccine was associated with fewer acute systemic symptoms at time of testing and fewer systemic and respiratory symptoms at Week 4, with some symptoms (fever, chills, loss of taste or smell, and sore throat) nearing resolution among vaccinated. These findings indicate a consistent additive benefit for BNT162b2 beyond prevention of severe disease, even with a new formulation and with the emergence of new sub-lineages.

Differently from our prior study, in this study we used PROMIS Fatigue 8a and noted that the mean fatigue T-score at Week 1 (60.2) was comparable to that of rheumatoid arthritis (8a T-score: 58.6) [27]. At the Week 4 data cutoff, fatigue levels did not return to levels similar to pre-COVID US. These findings highlight the impact of COVID-19 on fatigue levels, which appear comparable to the effects of chronic debilitating diseases during Week 1 of infection.

Additionally, during Week 1, the bivalent BA.4/5 BNT162b2 cohort reported less absenteeism and less impact on the workplace performance: participants vaccinated reported 18.8 workhours missed versus 25.4 among unvaccinated. Although not statistically significant, activity impairment scores were numerically lower in the vaccinated cohort across all time points.

Associations of bivalent BA.4/5 BNT162b2 with WPAI outcomes were more modest than those reported for the monovalent formulation, as they were observed during Week 1 only [4]. Also, we did not observe differences between the groups on HRQoL, differently from what observed for monovalent BNT162b2. Protection from COVID-19 bivalent vaccination has been observed to vary by Omicron sub-lineage, prior infection status, time since vaccination, time since prior infection, severity of infection and presence of risk factors [28,29]. Bivalent vaccination was targeted against BA.4/BA.5 Omicron sub-lineages and, while real-world evidence studies suggested cross-protection against successive sub-lineages, XBB was reported to be highly immune-evasive, triggering the need for a new, more closely matched, XBB vaccine formulation for 2023 / 2024 campaigns [30].

Considering that time since vaccination was relatively similar between the two studies (5-6 months), the more modest associations could be explained by the potential for reduced and less durable protection of the bivalent vaccine against XBB. In addition, although we controlled for prior infection, we could not measure time since prior infection. Accordingly, we could not sufficiently assess levels of immunity. Evidence has shown that hybrid immunity correlates with higher level of antibodies, improved protection against severe COVID-19 disease and potentially longer duration of protection, all of which could have impacted time of vaccination and our estimates [31].

This study has several strengths compared with published research. It is one of a limited number assessing diverse PROs associated with COVID-19 at community pharmacies. As such, it adds to existing evidence to contribute a holistic picture of humanistic outcomes associated with symptomatic COVID-19, assessed directly from a patient’s perspective. From an internal validity perspective, the study enrolled patients within days from testing positive and prospectively collected survey-based data shortly after infection, potentially minimizing recall bias. Moreover, the study leveraged widely used validated PRO instruments (EQ-5D-5L, WPAI, PROMIS Fatigue) and a questionnaire capturing a comprehensive symptoms list aligned to CDC research. With asymptomatic infections excluded by design, our estimates can be interpreted exclusively as related to symptomatic disease, potentially reducing the risk of overestimating the prevalence of acute COVID-19 symptoms among symptomatic. Further, both model-based and matching analyses yielded consistent results. Finally, with all study activities carried out virtually, this study piloted an innovative approach to agile and digitally enabled research during a pandemic.

The study is subject to several limitations. As previously described [5], all data collected were self-reported, subject to missingness, errors, recall bias, social desirability bias and selection bias associated with survey drop-off. Out of 643 participants, 26.7% were lost to follow-up at Week 4, possibly due to survey fatigue. Such drop-off rate should be interpreted in the context of participants being asked not to skip surveys. Such a strict requirement allowed for a clean assessment of changes in outcomes prevalence over time, but at the cost of attrition. Other limitations include over-representation of females, the relatively healthy baseline status of the population, exclusion of pediatrics, the fact that the study did not assess symptom severity, immunity levels, and that WPAI analyses were impacted by smaller sample size. Moreover, despite adjusting for several covariates in the model, risk of residual confounding may remain.

These findings may not be generalizable to other settings, prior or future variants, other countries, time periods and populations that were excluded. Finally, this study did not explore patient views, perceptions, and barriers to prevention.

While this study contributes to addressing knowledge gaps on symptomatology and PROs associated with COVID-19, symptoms are numerous, heterogeneous, and characterization of the acute infection continues to evolve. To our knowledge, this is the first study that provides estimates of the impact of bivalent BA.4/5 BNT162b2 on PROs in the context of high seroprevalence. As such, it adds novel insights and, when taken together with the prior monovalent BNT162b2 study^4,5^ and similar studies^1-3^, it solidifies evidence indicating that the effectiveness of BA.4/5 BNT162b2 on COVID-19 disease could translate to extra benefits of reduction in the frequency and burden of symptoms, supporting faster recovery and return to work. Future studies, especially evaluating variant-updated COVID-19 vaccine formulations, could corroborate these findings with different designs (for example, with a test-negative control), use of COVID-19 specific validated instruments, and assess these outcomes in subgroups defined by socio-demographic characteristics and antiviral treatment history.

## CONCLUSION

This study shows that COVID-19 adversely affects patients’ well-being, productivity and activity levels. Additionally, the results show the benefits of bivalent BA.4/5 BNT162b2 vaccine in alleviating symptoms and enhancing work productivity after acute SARS-CoV2 infection. These findings, taken together with existing evidence, show a consistent additive benefit of BNT162b2 beyond traditional endpoints, even with a new formulation and evolving sub-lineages.

## Author Contributions

All named authors meet the International Committee of Medical Journal Editors (ICMJE) criteria for authorship for this article.

Conceptualization, M.D.F., J.C.C., L.A.T., H.C., K.E.A., L.P., and X.S.;

Methodology, M.D.F., J.C.C., A.Y., K.E.A., L.P, S.M.C.L., and X.S.;

Formal Analysis, X.S.

Investigation, L.A.T. and X.S.;

Resources, M.D.F., J.C.C., L.A.T., A.Y., M.B.A., K.E.A., T.M.P., L.P., and S.M.C.L.;

Data Curation, X.S.

Writing—Original Draft Preparation, M.D.F., J.C.C., L.A.T., A.Y., K.E.A., T.M.P., L.P., S.M.C.L., and X.S.;

Writing—Review & Editing, M.D.F., J.C.C., L.A.T., H.C., A.Y., M.B.A., K.A., T.M.P., L.P., S.M.C.L., and X.S.;

Supervision, M.D.F., J.C.C., H.C., M.B.A., and L.P.;

Project Administration, M.D.F., L.A.T., T.M.P.; Funding Acquisition, M.D.F. and T.M.P..

## Institutional Review Board Statement

The study was conducted according to the guidelines of the Declaration of Helsinki and approved by the Sterling Institutional Review Board (C4591034 approved 13-Jan-2022).

## Informed Consent Statement

Informed consent was electronically obtained from all participants involved in the study via the CVS Health E-Consent platform. Participants were informed of their right to refuse or withdraw from the study at any time. No participants can be individually identified from the contents of this paper, therefore no additional individual participant consent for publication was required.

## Data Availability Statement

Aggregated data that support the findings of this study are available upon reasonable request from the corresponding author M.D.F., subject to review. These data are not publicly available due to them containing information that could compromise research participant privacy/consent.

## Acknowledgements

The authors acknowledge Meghan Gavaghan and Nancy Gifford (Pfizer employees), as well as Joseph Ferenchick, Shiyu Lin, Alexandra Berk and Shawn Edmonds (CVS Health employees) for specific contributions to this research project. Editorial support was provided by Leena Samuel at CVS Health and was funded by Pfizer.

