## Supplementary tables and figures for "Impact of bivalent BA.4/5 BNT162b2 COVID-19 vaccine on acute symptoms, quality of life, work productivity and activity levels among symptomatic US adults testing positive for SARS-CoV-2 at a national retail pharmacy": Vaccines_Bivalent-PROs_Supplemental material.docx

**Supplementary Material**

Supplemental Table 1 Patient Characteristics by Enrollment Status

Supplemental Table 2 Patient Characteristics by Vaccination Status after matching

Supplemental Table 3: Trajectory of acute COVID-19 symptoms at time of testing, Week 1, Week 2 and Week 4 after matching

Supplemental Table 4 Mixed Models for Repeated Measurements of EQ-5D-5L and WPAI-GH scores

**Supplemental Table 5.** EQ-5D-5L and WPAI-GH Scores post-matching

Supplemental Figure 1 SARS-CoV-2 Variants proportions in the US

Supplemental Figure 2 Prevalence of acute COVID-19 symptoms by vaccination status

Supplemental Table 1 Patient Characteristics by Enrollment Status

|  | | All | Included | Excluded | *P* ^a^ |
| --- | --- | --- | --- | --- | --- |
| Total, n (%) | | 21,027 | 643 | 20,384 |  |
| Age, years | |  |  |  |  |
| Mean, SD | | 45.3 (16.9) | 46.5 (15.9) | 45.2 (17.0) | 0.064 |
| Age group, n (%) | |  |  |  | 0.001 |
| 18-29 | | 4,749 (22.6) | 109 (17.0) | 4,640 (22.8) |  |
| 30-49 | | 7,661 (36.4) | 257 (40.0) | 7,404 (36.3) |  |
| 50-64 | | 5,276 (25.1) | 167 (26.0) | 5,109 (25.1) |  |
| 65-74 | | 2,371 (11.3) | 89 (13.8) | 2,282 (11.2) |  |
| ≥75 | | 970 (4.6) | 21 (3.2) | 949 (4.7) |  |
| Gender, n (%) | |  |  |  | <0.001 |
| Female | | 12,441 (59.2) | 452 (70.3) | 11,989 (58.8) |  |
| Male | | 8,428 (40.1) | 185 (28.8) | 8,243 (40.4) |  |
| Unknown | | 158 (0.7) | 6 (0.9) | 152 (0.8) |  |
| Race / Ethnicity, n (%) | |  |  |  | <0.001 |
| White or Caucasian (not Hispanic or Latino) | | 8,626 (41.0) | 374 (58.2) | 8,252 (40.5) |  |
| Black or African American | | 3,014 (14.3) | 57 (8.9) | 2,957 (14.5) |  |
| Hispanic | | 5,094 (24.2) | 99 (15.4) | 4,995 (24.5) |  |
| Asian | | 2,462 (11.7) | 63 (9.8) | 2,399 (11.8) |  |
| Patient Refused | | 1,051 (5.0) | 22 (3.4) | 1,029 (5.1) |  |
| Other | | 777 (3.7) | 28 (4.3) | 749 (3.7) |  |
| Missing | | 3 (0.0) | 0 (0.0) | 3 (0.0) |  |
| US Geographic Region, n (%) | |  |  |  | 0.037 |
| Northeast | | 2,493 (11.9) | 88 (13.7) | 2,405 (11.8) |  |
| South | | 9,517 (45.3) | 260 (40.4) | 9,257 (45.4) |  |
| Midwest | | 3,847 (18.3) | 141 (21.9) | 3,706 (18.2) |  |
| West | | 5,168 (24.6) | 154 (24.0) | 5,014 (24.6) |  |
| Missing | | 2 (0.0) | 0 (0.0) | 2 (0.0) |  |
| CMS Geographic Region (n, %) | |  |  |  | 0.002 |
| Region 1: ME, NH, VT, MA, CT, RI | | 1,088 (5.2) | 43 (6.7) | 1,045 (5.1) |  |
| Region 2: NY, NJ, PR, VI | | 684 (3.3) | 21 (3.3) | 663 (3.3) |  |
| Region 3: PA, DE, MD, DC, WV, VA | | 1,611 (7.7) | 52 (8.1) | 1,559 (7.7) |  |
| Region 4: KY, TN, NC, SC, GA, MS, AL, FL | | 4,742 (22.6) | 146 (22.7) | 4,596 (22.5) |  |
| Region 5: MN, WI, IL, MI, IN, OH | | 3,507 (16.7) | 132 (20.5) | 3,375 (16.6) |  |
| Region 6: NM, OK, AR, TX, LA | | 3,967 (18.9) | 88 (13.7) | 3,879 (19.0) |  |
| Region 7: NE, IA, KS, MO | | 336 (1.6) | 9 (1.4) | 327 (1.6) |  |
| Region 8: MT, ND, SD, WY, UT, CO | | 60 (0.3) | 5 (0.8) | 55 (0.3) |  |
| Region 9: CA, NV, AZ, GU | | 4,958 (23.6) | 142 (22.1) | 4,816 (23.6) |  |
| Region 10: AK, WA, OR, ID | | 73 (0.4) | 5 (0.8) | 68 (0.3) |  |
| Missing | | 1 (0.0 | 0 (0.0) | 1 (0.0) |  |
| Social vulnerability index, Mean (SD)^b^ | | 0.48 (0.2) | 0.45 (0.2) | 0.48 (0.2) | <0.001 |
| Previously Tested Positive, n (%) | |  |  |  | 0.532 |
| No | | 10,607 (50.4) | 338 (52.6) | 10,269 (50.4) |  |
| Yes | | 8,844 (42.1) | 268 (41.7) | 8,576 (42.1) |  |
| Missing | | 1,576 (7.5) | 37 (5.8) | 1,539 (7.6) |  |
| Number of comorbidities, Mean (SD) | | 0.30 (0.7) | 0.40 (0.8) | 0.30 (0.7) | 0.001 |
| At least 1 comorbidity, n (%) | | 4,138 (19.7) | 165 (25.7) | 3,973 (19.5) | <0.001 |
| Self-Reported Comorbidity, n (%) | |  |  |  |  |
| Asthma or Chronic Lung Disease | | 738 (3.5) | 33 (5.1) | 705 (3.5) | 0.023 |
| Cirrhosis of the liver | | 51 (0.2) | 2 (0.3) | 49 (0.2) | 0.720 |
| Immunocompromised Conditions or Weakened Immune System^c^ | | 106 (0.5) | 2 (0.3) | 104 (0.5) | 0.483 |
| Diabetes | | 1,277 (6.1) | 39 (6.1) | 1,238 (6.1) | 0.993 |
| Heart Conditions or Hypertension | | 2,783 (13.2) | 104 (16.2) | 2,679 (13.1) | 0.026 |
| Overweight or obesity | | 1,625 (7.7) | 80 (12.4) | 1,545 (7.6) | <0.001 |
| **Index day^d^ acute COVID-19 symptoms** | |  |  |  |  |
| Number of symptoms, Mean, SD | 5.30 (2.40) | | 5.30 (2.30) | 5.30 (2.4) | 0.484 |
| Fever | 9,033 (43.0) | | 268 (41.7) | 8,765 (43.0) | 0.506 |
| Chills | 3,225 (15.3) | | 101 (15.7) | 3,124 (15.3) | 0.791 |
| Muscle or Body Aches | 16,798 (79.9) | | 491 (76.4) | 16,307 (80.0) | 0.023 |
| Headache | 11,058 (52.6) | | 332 (51.6) | 10,726 (52.6) | 0.622 |
| Fatigue | 1,019 (4.9) | | 25 (3.9) | 994 (4.9) | 0.251 |
| Shortness of Breath or Difficulty Breathing | 9,956 (47.3) | | 297 (46.2) | 9,659 (47.4) | 0.550 |
| Cough | 11,935 (56.8) | | 390 (60.7) | 11,545 (56.6) | 0.043 |
| Sore Throat | 2,879 (13.7) | | 89 (13.8) | 2,790 (13.7) | 0.911 |
| New/Recent Loss of Taste or Smell | 12,441 (59.2) | | 424 (65.9) | 12,017 (59.0) | 0.000 |
| Congestion or Runny Nose | 13,100 (62.3) | | 417 (64.9) | 12,683 (62.2) | 0.175 |
| Nausea or Vomiting | 16,398 (78.0) | | 525 (81.6) | 15,873 (77.9) | 0.023 |
| Diarrhea | 3,230 (15.4) | | 80 (12.4) | 3,150 (15.5) | 0.037 |

CMS: Centers for Medicare and Medicaid Services; SD: Standard Deviation

^a^ *P* value refers to the comparison between Included and Excluded.

^b^ SVI ranges from 0 to 1. A community with higher value is more socially vulnerable.

^c^ Immunocompromised conditions includes compromised immune system (such as from immuno-compromising drugs, solid organ or blood stem cell transplant, HIV, or other conditions), conditions that result in a weakened immune system, including kidney failure or end stage renal disease

^d^ COVID-19 test nasal swab day

Supplemental Table 2 Patient Characteristics by Vaccination Status after matching

|  | All | BNT162b2 | Unvaccinated | *P* ^a^ |
| --- | --- | --- | --- | --- |
| Total, n (%) | 643 | 316 (49.1) | 327 (50.9) |  |
| Age, years |  |  |  |  |
| Mean, SD | 46.5 (15.9) | 46.6 (15.9) | 46.4 (15.9) | 0.917 |
| Age group, n (%) |  |  |  | 0.776 |
| 18-29 | 109 (17.0) | 54 (17.0) | 56 (17.0) |  |
| 30-49 | 257 (40.0) | 126 (40.0) | 131 (40.0) |  |
| 50-64 | 167 (26.0) | 82 (26.0) | 85 (26.0) |  |
| 65-74 | 89 (13.8) | 44 (13.8) | 45 (13.8) |  |
| ≥75 | 21 (3.2) | 10 (3.2) | 11 (3.2) |  |
| Gender, n (%) |  |  |  | 1.000 |
| Female | 458 (71.1) | 221 (70.0) | 236 (72.2) |  |
| Male | 178 (27.7) | 91 (28.6) | 88 (26.8) |  |
| Unknown | 7 (1.2) | 4 (1.3) | 3.1 (1.0) |  |
| Race / Ethnicity, n (%) |  |  |  | 1.000 |
| White or Caucasian (not Hispanic or Latino) | 374 (58.2) | 184 (58.2) | 190 (58.2) |  |
| Black or African American | 57 (8.9) | 28 (8.9) | 29 (8.9) |  |
| Hispanic | 99 (15.4) | 49 (15.4) | 50 (15.4) |  |
| Asian | 63 (9.8) | 31 (9.8) | 32 (9.8) |  |
| Patient Refused | 22 (3.4) | 11 (3.4) | 11 (3.4) |  |
| Other | 28 (4.3) | 14 (4.3) | 14 (4.3) |  |
| US Geographic Region, n (%) |  |  |  | 0.114 |
| Northeast | 88 (13.7) | 43 (13.7) | 45 (13.7) |  |
| South | 260 (40.4) | 128 (40.4) | 132 (40.4) |  |
| Midwest | 141 (21.9) | 69 (21.9) | 72 (21.9) |  |
| West | 154 (24.0) | 76 (24.0) | 79 (24.0) |  |
| CMS Geographic Region (n, %) |  |  |  | 0.090 |
| Region 1: ME, NH, VT, MA, CT, RI | 43 (6.8) | 23 (7.2) | 21 (6.4) |  |
| Region 2: NY, NJ, PR, VI | 23 (3.6) | 10 (3.3) | 13 (3.9) |  |
| Region 3: PA, DE, MD, DC, WV, VA | 57 (8.8) | 34 (10.9) | 22 (6.7) |  |
| Region 4: KY, TN, NC, SC, GA, MS, AL, FL | 143 (22.3) | 69 (21.9) | 74 (22.7) |  |
| Region 5: MN, WI, IL, MI, IN, OH | 132 (20.5) | 64 (20.3) | 68 (20.7) |  |
| Region 6: NM, OK, AR, TX, LA | 83 (12.8) | 35 (11.0) | 48 (14.6) |  |
| Region 7: NE, IA, KS, MO | 9 (1.4) | 5 (1.6) | 4 (1.2) |  |
| Region 8: MT, ND, SD, WY, UT, CO | 5 (0.7) | 4 (1.2) | 1 (0.2) |  |
| Region 9: CA, NV, AZ, GU | 143 (22.2) | 66 (20.7) | 77 (23.6) |  |
| Region 10: AK, WA, OR, ID | 6 (0.9) | 6 (1.9) | 0 (0.0) |  |
| Social vulnerability index, Mean (SD)^b^ | 0.44 (0.2) | 0.44 (0.2) | 0.45 (0.2) | 0.801 |
| Previously Tested Positive, n (%) |  |  |  | 0.090 |
| No | 328 (54.6) | 170 (58.1) | 158 (51.2) |  |
| Yes | 273 (45.4) | 123 (41.9) | 151 (48.8) |  |
| Missing | 37 | 20 | 17 |  |
| Number of comorbidities, Mean (SD) | 0.40 (0.8) | 0.40 (0.8) | 0.40 (0.8) | 0.770 |
| At least 1 comorbidity, n (%) | 165 (25.7) | 81 (25.7) | 84 (25.7) | 1.000 |
| Self-Reported Comorbidity, n (%) |  |  |  |  |
| Asthma or Chronic Lung Disease | 37 (5.8) | 20 (6.4) | 17 (5.2) | 0.506 |
| Cirrhosis of the liver | 2 (0.4) | 0 (0.0) | 2 (0.7) | 0.133 |
| Immunocompromised Conditions or Weakened Immune System^c^ | 2 (0.3) | 1 (0.3) | 1 (0.2) | 0.849 |
| Diabetes | 39 (6.0) | 21 (6.5) | 18 (5.5) | 0.596 |
| Heart Conditions or Hypertension | 100 (15.5) | 44 (14.0) | 56 (17.0) | 0.292 |
| Overweight or obesity | 80 (12.4) | 39 (12.2) | 41 (12.6) | 0.878 |
| Smoking | 25 (3.8) | 11 (3.4) | 14 (4.2) | 0.572 |
| Paxlovid prescription, n (%) |  |  |  | 1.000 |
| No | 495 (77.0) | 243 (77.0) | 252 (77.0) |  |
| Yes | 148 (23.0) | 73 (23.0) | 75 (23.0) |  |
| Missing | 2 | 2 | 0 |  |

CMS: Centers for Medicare and Medicaid Services; SD: Standard Deviation

^a^ *P* value refers to the comparison between BNT162b2 and Unvaccinated.

^b^ SVI ranges from 0 to 1. A community with higher value is more socially vulnerable.

^c^ Immunocompromised conditions includes compromised immune system (such as from immuno-compromising drugs, solid organ or blood stem cell transplant, HIV, or other conditions), conditions that result in a weakened immune system, including cancer treatment, and kidney failure or end stage renal disease

Supplemental Table 3 Trajectory of acute COVID-19 symptoms at time of testing, Week 1, Week 2 and Week 4 after matching

|  | All | BNT162b2 | Unvaccinated | *P* ^a^ |
| --- | --- | --- | --- | --- |
| **Index day (time of testing)** |  |  |  |  |
| n | 643 | 316 (49.1) | 327 (50.9) |  |
| Mean number of symptoms (SD) | 5.3 (2.3) | 5.1 (2.4) | 5.5 (2.2) | 0.008 |
| Median (Q1, Q3) | 5 (3.0, 7.0) | 5 (3.0, 7.0) | 6 (4.0, 7.0) |  |
| Min, max | 1, 12 | 1, 12 | 1, 11 |  |
| Missing | 0 | 0 | 0 |  |
| Number of ARI symptoms, n (%) |  |  |  | 0.022 |
| <3 | 79 (12.4) | 46 (14.6) | 33 (10.1) |  |
| 3-5 | 273 (42.5) | 143 (45.2) | 130 (39.8) |  |
| 6-8 | 235 (36.5) | 97 (30.8) | 137 (42.0) |  |
| 9+ | 56 (8.7) | 30 (9.4) | 27 (8.1) |  |
| Systemic symptoms, n (%) | 569 (88.4) | 271 (85.7) | 298 (91.1) | 0.032 |
| Fever | 263 (40.8) | 117 (37.0) | 146 (44.6) | 0.050 |
| Chills | 327 (50.8) | 142 (44.8) | 185 (56.6) | 0.003 |
| Muscle or Body Aches | 285 (44.2) | 120 (37.8) | 165 (50.4) | 0.002 |
| Headache | 417 (64.8) | 195 (61.5) | 222 (68.0) | 0.085 |
| Fatigue | 417 (64.9) | 201 (63.7) | 216 (66.1) | 0.523 |
| Respiratory symptoms, n (%) | 629 (97.8) | 311 (98.3) | 318 (97.3) | 0.375 |
| Shortness of Breath or Difficulty Breathing | 100 (15.5) | 50 (15.7) | 50 (15.4) | 0.910 |
| Cough | 493 (76.7) | 237 (74.9) | 256 (78.4) | 0.304 |
| Sore Throat | 393 (61.1) | 196 (61.9) | 197 (60.3) | 0.685 |
| New/Recent Loss of Taste or Smell | 83 (12.9) | 38 (12.1) | 45 (13.7) | 0.544 |
| Congestion or Runny Nose | 531 (82.5) | 260 (82.3) | 271 (82.8) | 0.862 |
| GI symptoms, n (%) | 88 (13.6) | 37 (11.6) | 51 (15.6) | 0.143 |
| Nausea or Vomiting | 23 (3.6) | 10 (3.3) | 12 (3.8) | 0.724 |
| Diarrhea | 76 (11.8) | 33 (10.3) | 43 (13.2) | 0.264 |
| **Week 1** |  |  |  |  |
| n | 566 | 285 (50.4) | 281 (49.6) |  |
| Mean number of symptoms (SD) | 2.5 (1.7) | 2.5 (1.7) | 2.6 (1.8) | 0.365 |
| Median, Q1-Q3 | 2 (1.0, 3.0) | 2 (1.0, 3.0) | 2 (1.0, 3.0) |  |
| Min, max | 1, 10 | 1, 10 | 1, 9 |  |
| Missing | 1 | 0 | 1 |  |
| Number of ARI symptoms |  |  |  | 0.466 |
| <3 | 350 (61.7) | 183 (64.0) | 167 (59.3) |  |
| 3-5 | 184 (32.4) | 90 (31.5) | 94 (33.4) |  |
| 6-8 | 24 (4.3) | 9 (3.2) | 15 (5.4) |  |
| 9+ | 9 (1.6) | 4 (1.3) | 5 (1.8) |  |
| Missing | 1 (0.1) | 0 (0.0) | 1 (0.2) |  |
| Systemic symptoms, n (%) | 329 (58.1) | 155 (54.5) | 174 (61.7) | 0.083 |
| Fever | 15 (2.6) | 3 (1.2) | 12 (4.1) | 0.016 |
| Chills | 20 (3.5) | 9 (3.3) | 11 (3.8) | 0.607 |
| Muscle or Body Aches | 83 (14.7) | 33 (11.7) | 50 (17.7) | 0.034 |
| Headache | 123 (21.8) | 52 (18.4) | 71 (25.2) | 0.042 |
| Fatigue | 281 (49.6) | 136 (47.7) | 145 (51.6) | 0.337 |
| Respiratory symptoms, n (%) | 415 (73.2) | 209 (73.5) | 206 (73.0) | 0.720 |
| Shortness of Breath or Difficulty Breathing | 89 (15.7) | 45 (15.9) | 42 (15.4) | 0.820 |
| Cough | 276 (48.7) | 137 (48.0) | 139 (49.4) | 0.816 |
| Sore Throat | 79 (13.9) | 42 (14.8) | 37 (13.0) | 0.591 |
| New/Recent Loss of Taste or Smell | 73 (12.9) | 36 (12.7) | 37 (13.2) | 0.741 |
| Congestion or Runny Nose | 249 (43.9) | 134 (47.0) | 115 (40.7) | 0.130 |
| GI symptoms, n (%) | 41 (7.3) | 23 (7.9) | 19 (6.7) | 0.643 |
| Nausea or Vomiting | 2 (0.4) | 1 (0.3) | 1 (0.5) | 0.741 |
| Diarrhea | 39 (6.9) | 22 (7.6) | 18 (6.3) | 0.583 |
| **Week 2** |  |  |  |  |
| n | 530 | 269 (50.8) | 261 (49.2) |  |
| Mean number of symptoms (SD) | 1.9 (1.3) | 1.9 (1.3) | 2.0 (1.4) | 0.182 |
| Median, Q1-Q3 | 1 (1.0, 2.0) | 1 (1.0, 2.0) | 1 (1.0, 3.0) |  |
| Min, max | 1, 8 | 1, 8 | 1, 8 |  |
| Missing | 0 | 0 | 0 |  |
| Number of ARI symptoms |  |  |  | 0.242 |
| <3 | 401 (75.7) | 210 (78.2) | 191 (73.1) |  |
| 3-5 | 117 (22.0) | 54 (19.9) | 63 (24.2) |  |
| 6-8 | 12 (2.3) | 5 (1.9) | 7 (2.7) |  |
| Systemic symptoms, n (%) | 254 (48.0) | 126 (47) | 128 (48.9) | 0.508 |
| Fever | 7 (1.4) | 3 (0.9) | 5 (1.8) | 0.405 |
| Chills | 13 (2.4) | 6 (2.1) | 7 (2.7) | 0.663 |
| Muscle or Body Aches | 72 (13.7) | 34 (12.5) | 39 (14.8) | 0.340 |
| Headache | 80 (15.2) | 34 (12.6) | 47 (17.8) | 0.084 |
| Fatigue | 218 (41.2) | 110 (40.7) | 109 (41.8) | 0.675 |
| Respiratory symptoms, n (%) | 270 (50.9) | 129 (47.8) | 141 (54.1) | 0.139 |
| Shortness of Breath or Difficulty Breathing | 48 (9.1) | 21 (7.9) | 27 (10.3) | 0.367 |
| Cough | 181 (34.1) | 91 (33.8) | 90 (34.4) | 0.853 |
| Sore Throat | 43 (8.2) | 16 (5.9) | 28 (10.5) | 0.039 |
| New/Recent Loss of Taste or Smell | 29 (5.4) | 16 (5.9) | 13 (4.8) | 0.870 |
| Congestion or Runny Nose | 128 (24.2) | 65 (24.1) | 64 (24.4) | 0.822 |
| GI symptoms, n (%) | 27 (5.1) | 13 (4.9) | 14 (5.3) | 0.855 |
| Nausea or Vomiting | 3 (0.6) | 3 (0.9) | 1 (0.3) | 0.302 |
| Diarrhea | 25 (4.7) | 12 (4.5) | 13 (5.0) | 0.784 |
| **Week 4** |  |  |  |  |
| n | 505 | 260 (51.5) | 245 (48.5) |  |
| Mean number of symptoms (SD) | 0.8 (1.3) | 0.7 (1.2) | 1.2 (1.0) | 0.004 |
| Median, Q1-Q3 | 0 (0.0, 1.0) | 0 (0.0, 1.0) | 1 (0.0, 2.0) |  |
| Min, max | 0, 8 | 0, 7 | 0, 8 |  |
| Missing | 1 | 0 | 1 |  |
| Number of ARI symptoms |  |  |  | 0.005 |
| 0 | 275 (54.4) | 161 (61.9) | 114 (46.5) |  |
| 1-2 | 182 (35.9) | 81 (31.0) | 101 (41.0) |  |
| 3-5 | 42 (8.3) | 15 (5.6) | 27 (11.0) |  |
| 6-8 | 7 (1.4) | 4 (1.5) | 3 (1.3) |  |
| Missing | 1 (0.1) | 0 (0.0) | 1 (0.2) |  |
| Systemic symptoms, n (%) | 161 (31.9) | 66 (25.5) | 95 (38.6) | 0.001 |
| Fever | 7 (1.4) | 3 (1.3) | 4 (1.4) | 0.889 |
| Chills | 7 (1.4) | 4 (1.7) | 3 (1.1) | 0.672 |
| Muscle or Body Aches | 39 (7.8) | 15 (5.8) | 24 (9.7) | 0.036 |
| Headache | 64 (12.6) | 29 (11.1) | 35 (14.2) | 0.324 |
| Fatigue | 115 (22.8) | 44 (17.1) | 71 (28.8) | 0.001 |
| Respiratory symptoms, n (%) | 129 (25.5) | 57 (22.1) | 72 (29.1) | 0.042 |
| Shortness of Breath or Difficulty Breathing | 48 (9.5) | 22 (8.6) | 26 (10.4) | 0.415 |
| Cough | 85 (16.8) | 39 (15.0) | 46 (18.8) | 0.162 |
| Sore Throat | 19 (3.7) | 5 (2.0) | 14 (5.5) | 0.042 |
| New/Recent Loss of Taste or Smell | 14 (2.8) | 6 (2.4) | 8 (3.3) | 0.405 |
| Congestion or Runny Nose | N/A | | | |
| GI symptoms, n (%) | 27 (5.3) | 12 (4.7) | 15 (6.0) | 0.537 |
| Nausea or Vomiting | 14 (2.8) | 5 (2.1) | 9 (3.5) | 0.348 |
| Diarrhea | 15 (2.9) | 7 (2.6) | 8 (3.2) | 0.694 |

^a^ *P* values refers to the comparison between BNT162b2 and Unvaccinated, after matching by time point on age, race/ethnicity, region, SVI category, Paxlovid use, ≥1 comorbidity.

Supplemental Table 4 Mixed Models for Repeated Measurements EQ-5D-5L and WPAI-GH Scores: Estimate (Standard Error)

|  | EQ VAS | EQ-5D-5L UI | Absenteeism | Presenteeism | Work Productivity Loss | Activity Impairment | Hours Missed due to health | Actual hours worked | PROMIS Fatigue 8(a) |
| --- | --- | --- | --- | --- | --- | --- | --- | --- | --- |
| Intercept | 25.0 (3.4) | 0.310 (0.043) | 3.6 (3.7) | 0.4 (4.8) | 6.3 (5.3) | 1.9 (3.8) | -0.2 (2.1) | 24.3 (2.8) | 27.1 (2.1) |
| Time |  |  |  |  |  |  |  |  |  |
| Day 3 / Week 1 | Ref | Ref | Ref | Ref | Ref | Ref | Ref | Ref | Ref |
| Week 2 | 11.5 (0.8) | 0.059 (0.009) | 56.5 (2.7) | 42.4 (2.7) | 53.5 (2.8) | 41.7 (2.1) | 21.9 (1.3) | -20.2 (1.3) | 12.8 (0.6) |
| Week 4 | 14.2 (0.9) | 0.077 (0.009) | 5.6 (2.0) | 6.8 (1.6) | 6.7 (2.0) | 5.8 (1.4) | 1.7 (1.1) | -2.2 (1.1) | 3.0 (0.5) |
| BNT162b2 |  |  |  |  |  |  |  |  |  |
| No | Ref | Ref | Ref | Ref | Ref | Ref | Ref | Ref | Ref |
| Yes | -0.8 (1.2) | -0.002 (0.012) | -0.9 (1.8) | 0.3 (2.4) | -0.9 (2.9) | -0.1 (2.0) | 0.3 (1.2) | -0.2 (1.4) | -0.2 (0.8) |
| BNT162b2by Time |  |  |  |  |  |  |  |  |  |
| Yes * Day 3/Week 1 | Ref | Ref | Ref | Ref | Ref | Ref | Ref | Ref | Ref |
| Yes * Week 2 | -0.2 (1.1) | 0.008 (0.013) | -11.3 (3.8) | -5.9 (3.8) | -5.1 (4.0) | -2.5 (2.9) | -6.4 (1.9) | 4.8 (1.9) | 0.0 (0.9) |
| Yes * Week 4 | 0.6 (1.3) | 0.004 (0.013) | 1.9 (2.9) | -2.0 (2.3) | 0.1 (2.9) | 0.7 (2.0) | 0.1 (1.5) | -0.7 (1.6) | 1.0 (0.7) |
| Baseline score | 0.6 (0.0) | 0.624 (0.039) | 0.0 (0.0) | 0.3 (0.0) | 0.2 (0.0) | 0.3 (0.0) | 0.1 (0.0) | 0.3 (0.0) | 0.3 (0.0) |
| Age group |  |  |  |  |  |  |  |  |  |
| 18-29 | Ref | Ref | Ref | Ref | Ref | Ref | Ref | Ref | Ref |
| 30-49 | -0.7 (1.1) | -0.010 (0.013) | -0.1 (2.1) | 6.0 (2.7) | 5.3 (3.0) | 4.7 (2.2) | 0.8 (1.1) | 2.6 (1.5) | 2.3 (0.9) |
| 50-64 | -1.6 (1.2) | -0.016 (0.015) | 3.3 (2.4) | 4.4 (3.2) | 5.0 (3.5) | 3.5 (2.5) | 1.7 (1.3) | 1.9 (1.7) | 2.0 (1.1) |
| 65-74 | 0.1 (1.4) | -0.005 (0.017) | 1.0 (3.8) | -2.9 (5.0) | -5.6 (5.5) | -1.4 (2.9) | -0.6 (2.1) | -3.3 (2.7) | 0.9 (1.2) |
| 75+ | -0.1 (2.3) | 0.007 (0.028) | -1.5 (9.5) | 9.2 (11.6) | 4.9 (12.5) | 0.2 (4.8) | -3.5 (4.5) | -0.6 (5.7) | 0.3 (2.0) |
| Gender |  |  |  |  |  |  |  |  |  |
| Female | Ref | Ref | Ref | Ref | Ref | Ref | Ref | Ref | Ref |
| Male | 1.6 (0.8) | 0.016 (0.010) | -0.2 (1.7) | -1.8 (2.3) | -0.9 (2.5) | -4.3 (1.7) | 1.3 (1.0) | 3.1 (1.2) | -3.1 (0.7) |
| Unknown | -5.2 (4.2) | -0.224 (0.050) | 3.6 (9.1) | 6.9 (13.2) | -1.6 (15.0) | 11.7 (8.7) | -0.1 (4.8) | -10.0 (6.0) | 7.5 (3.6) |
| Race/Ethnicity |  |  |  |  |  |  |  |  |  |
| White or Caucasian | Ref | Ref | Ref | Ref | Ref | Ref | Ref | Ref | Ref |
| Black or African American | 3.7 (1.3) | 0.028 (0.016) | 1.3 (2.9) | -2.5 (3.7) | -0.6 (4.0) | -2.1 (2.8) | 1.4 (1.5) | 0.9 (2.0) | -1.8 (1.2) |
| Hispanic | 1.1 (1.1) | -0.023 (0.014) | 3.5 (2.3) | 0.3 (3.0) | 2.3 (3.4) | 2.6 (2.4) | 1.4 (1.2) | -2.8 (1.6) | 0.6 (1.0) |
| Asian | 1.5 (1.3) | 0.022 (0.016) | -1.3 (2.6) | -5.9 (3.4) | -6.8 (3.8) | -4.0 (2.7) | 0.1 (1.5) | 0.1 (1.9) | -1.4 (1.1) |
| Patient Refused | 0.2 (2.1) | -0.030 (0.025) | 7.7 (4.3) | 0.5 (5.8) | 3.0 (6.6) | 8.3 (4.3) | 3.2 (2.3) | 2.4 (3.0) | 0.3 (1.8) |
| Other | 1.7 (1.8) | 0.000 (0.022) | -2.4 (3.8) | -9.4 (5.2) | -11.3 (5.6) | -1.3 (3.7) | -1.9 (2.1) | 3.5 (2.7) | -0.6 (1.6) |
| US Region |  |  |  |  |  |  |  |  |  |
| Northeast | Ref | Ref | Ref | Ref | Ref | Ref | Ref | Ref | Ref |
| South | 0.8 (1.2) | 0.026 (0.014) | -0.1 (2.4) | -0.7 (3.1) | -1.1 (3.4) | -1.7 (2.4) | -0.5 (1.3) | -0.1 (1.7) | -1.1 (1.0) |
| Midwest | -1.2 (1.2) | -0.002 (0.015) | -0.7 (2.5) | -1.1 (3.2) | -0.5 (3.5) | -0.7 (2.6) | -1.1 (1.3) | 0.9 (1.7) | -1.2 (1.1) |
| West | 0.9 (1.3) | 0.019 (0.015) | 0.5 (2.6) | -0.4 (3.3) | -0.1 (3.6) | -2.8 (2.6) | 0.5 (1.4) | 1.3 (1.8) | -1.1 (1.1) |
| Social Vulnerability Index |  |  |  |  |  |  |  |  |  |
| <0.25 | Ref | Ref | Ref | Ref | Ref | Ref | Ref | Ref | Ref |
| ≥0.25 and <0.5 | -1.0 (0.9) | -0.003 (0.012) | 1.3 (2.0) | -1.1 (2.6) | -0.6 (2.8) | 0.7 (1.9) | 1.3 (1.1) | -1.9 (1.4) | 0.0 (0.8) |
| ≥0.5 and <0.75 | -0.3 (1.1) | -0.008 (0.013) | 0.0 (2.2) | -1.3 (2.9) | -2.0 (3.2) | -0.2 (2.2) | 0.0 (1.2) | -1.9 (1.6) | -0.3 (0.9) |
| ≥0.75 | -1.0 (1.3) | -0.018 (0.016) | 0.9 (2.8) | -3.6 (3.6) | -3.1 (4.0) | -1.3 (2.7) | 0.5 (1.5) | -0.9 (2.0) | 0.6 (1.1) |
| ≥1 Comorbidity | -0.9 (0.9) | -0.013 (0.010) | -0.4 (1.8) | 3.2 (2.3) | 2.9 (2.5) | 2.2 (1.8) | 0.9 (1.0) | -1.5 (1.2) | 0.2 (0.7) |
| Previously tested positive | 1.2 (0.7) | 0.006 (0.009) | -0.7 (1.5) | -3.0 (1.9) | -4.7 (2.1) | -3.6 (1.5) | -0.7 (0.8) | -0.8 (1.0) | -1.2 (0.6) |
| Number of Acute COVID Symptoms on index day | -0.7 (0.2) | -0.013 (0.002) | 0.1 (0.3) | 1.9 (0.4) | 1.8 (0.5) | 1.9 (0.3) | 0.1 (0.2) | 0.2 (0.2) | 1.1 (0.1) |
| Paxlovid prescription | -1.1 (0.9) | -0.016 (0.011) | 0.1 (1.9) | 0.5 (2.4) | 0.8 (2.7) | 4.2 (1.8) | 1.1 (1.0) | 1.0 (1.3) | 2.0 (0.8) |

**Supplemental Table 5.** EQ-5D-5L and WPAI-GH Scores post-matching

|  | BNT162b2 | | | | | | | Unvaccinated | | | | | | | Difference in Change from Baseline Between Cohorts | | |
| --- | --- | --- | --- | --- | --- | --- | --- | --- | --- | --- | --- | --- | --- | --- | --- | --- | --- |
|  | Score | | Change from Baseline ^b^ | | | | | Score | | Change from Baseline | | | | |  |  |  |
|  | n | Mean (SD) | n | Mean (SD) | *P* ^c^ | | ES_w_^d^ | n | Mean (SD) | n | | Mean (SD) | *P* ^c^ | ES_w_^d^ | Mean (SD) | *P* ^e^ | ES_b_^f^ |
| EQ VAS |  |  |  |  |  |  | |  |  |  |  | |  |  |  |  |  |
| Baseline | 314 | 85.4 (10.8) |  |  |  |  | | 326 | 86.4 (12.5) |  |  | |  |  | -0.9 (11.7) | 0.307 | -0.08 |
| Day 3 | 314 | 70.1 (16.4) | 314 | -15.3 (13.7) | 0.000 | -1.12 | | 324 | 71.5 (17.0) | 324 | -14.9 (15.1) | | 0.000 | -0.99 | -0.4 (14.4) | 0.731 | -0.03 |
| Week 2 | 266 | 81.7 (12.2) | 265 | -3.6 (10.8) | 0.000 | -0.33 | | 258 | 83.1 (11.7) | 258 | -3.7 (10.5) | | 0.000 | -0.35 | 0.1 (10.7) | 0.921 | 0.01 |
| Week 4 | 258 | 85.2 (11.0) | 257 | -0.2 (10.3) | 0.777 | -0.02 | | 240 | 85.6 (11.0) | 240 | -1.2 (11.5) | | 0.122 | -0.10 | 1.0 (10.9) | 0.322 | 0.09 |
| Utility Index |  |  |  |  |  |  | |  |  |  |  | |  |  |  |  |  |
| Baseline^e^ | 316 | 0.926 (0.116) |  |  |  |  | | 327 | 0.928 (0.104) |  |  | |  |  | -0.002 (0.110) | 0.840 | -0.02 |
| Day 3 | 316 | 0.816 (0.188) | 316 | -0.110 (0.155) | 0.000 | -0.71 | | 327 | 0.820 (0.162) | 327 | -0.108 (0.152) | | 0.000 | -0.71 | -0.002 (0.154) | 0.879 | -0.01 |
| Week 2 | 269 | 0.891 (0.140) | 269 | -0.036 (0.119) | 0.000 | -0.31 | | 261 | 0.883 (0.140) | 261 | -0.050 (0.138) | | 0.000 | -0.37 | 0.014 (0.129) | 0.212 | 0.11 |
| Week 4 | 260 | 0.904 (0.137) | 260 | -0.022 (0.125) | 0.004 | -0.18 | | 246 | 0.898 (0.133) | 246 | -0.038 (0.126) | | 0.000 | -0.30 | 0.016 (0.125) | 0.161 | 0.12 |
| Absenteeism |  |  |  |  |  |  | |  |  |  |  | |  |  |  |  |  |
| Baseline^e^ | 182 | 7.3 (20.7) |  |  |  |  | | 205 | 11.4 (25.1) |  |  | |  |  | -4.1 (23.1) | 0.084 | -0.18 |
| Week 1 | 196 | 51.0 (37.3) | 178 | 42.8 (39.6) | 0.000 | 1.08 | | 224 | 62.8 (33.8) | 200 | 51.3 (40.1) | | 0.000 | 1.28 | -8.5 (39.9) | 0.039 | -0.21 |
| Week 2 | 182 | 11.6 (23.8) | 168 | 4.8 (29.3) | 0.036 | 0.16 | | 202 | 12.5 (24.0) | 183 | 1.7 (33.5) | | 0.488 | 0.05 | 3.1 (31.6) | 0.367 | 0.10 |
| Week 4 | 174 | 5.3 (15.2) | 159 | -2.5 (24.6) | 0.201 | -0.10 | | 188 | 6.4 (15.1) | 167 | -4.8 (27.0) | | 0.022 | -0.18 | 2.3 (25.9) | 0.420 | 0.09 |
| Presenteeism |  |  |  |  |  |  | |  |  |  |  | |  |  |  |  |  |
| Baseline^e^ | 179 | 12.0 (20.5) |  |  |  |  | | 197 | 15.4 (25.2) |  |  | |  |  | -3.3 (23.1) | 0.163 | -0.14 |
| Week 1 | 155 | 49.1 (30.2) | 143 | 37.7 (29.3) | 0.000 | 1.29 | | 155 | 56.5 (30.2) | 137 | 42.9 (30.9) | | 0.000 | 1.39 | -5.2 (30.1) | 0.150 | -0.17 |
| Week 2 | 174 | 20.0 (23.3) | 159 | 7.6 (26.0) | 0.000 | 0.29 | | 196 | 21.5 (23.9) | 172 | 8.2 (29.1) | | 0.000 | 0.28 | -0.5 (27.7) | 0.865 | -0.02 |
| Week 4 | 174 | 13.3 (21.7) | 156 | 2.5 (26.7) | 0.236 | 0.10 | | 188 | 16.1 (20.5) | 161 | 1.6 (27.0) | | 0.447 | 0.06 | 0.9 (26.8) | 0.761 | 0.03 |
| Work productivity loss | | | | | | | | | | | | | | | | | |
| Baseline^e^ | 179 | 16.0 (24.4) |  |  |  |  | | 197 | 19.3 (28.7) |  |  | |  |  | -3.3 (26.7) | 0.231 | -0.12 |
| Week 1 | 155 | 65.0 (28.5) | 143 | 48.9 (32.9) | 0.000 | 1.49 | | 155 | 73.7 (26.5) | 137 | 54.6 (33.0) | | 0.000 | 1.65 | -5.7 (33.0) | 0.151 | -0.17 |
| Week 2 | 174 | 25.2 (27.6) | 159 | 8.6 (31.8) | 0.001 | 0.27 | | 196 | 27.2 (27.9) | 172 | 9.8 (35.7) | | 0.000 | 0.27 | -1.2 (33.9) | 0.753 | -0.03 |
| Week 4 | 174 | 16.9 (25.5) | 156 | 2.0 (31.1) | 0.411 | 0.07 | | 188 | 20.1 (24.4) | 161 | 2.0 (32.4) | | 0.439 | 0.06 | 0.1 (31.7) | 0.985 | 0.00 |
| Activity impairment | | | | | | | | | | | | | | | | | |
| Baseline^e^ | 284 | 14.0 (22.0) |  |  |  |  | | 281 | 18.5 (27.8) |  |  | |  |  | -4.5 (25.0) | 0.033 | -0.18 |
| Week 1 | 282 | 55.5 (29.2) | 281 | 41.7 (33.3) | 0.000 | 1.25 | | 276 | 59.6 (29.0) | 275 | 40.6 (35.5) | | 0.000 | 1.15 | 1.0 (34.4) | 0.723 | 0.03 |
| Week 2 | 269 | 22.8 (24.6) | 268 | 8.4 (26.9) | 0.000 | 0.31 | | 261 | 24.4 (25.8) | 260 | 6.6 (29.0) | | 0.000 | 0.23 | 1.8 (27.9) | 0.451 | 0.07 |
| Week 4 | 260 | 15.3 (21.2) | 259 | 0.8 (26.2) | 0.623 | 0.03 | | 245 | 18.7 (23.9) | 244 | 1.0 (27.6) | | 0.580 | 0.04 | -0.2 (26.9) | 0.941 | -0.01 |
| Hours missed due to health | | | | | | | | | | | | | | | | | |
| Baseline^e^ | 192 | 3.4 (10.2) |  |  |  |  | | 216 | 6.1 (14.4) |  |  | |  |  | -2.8 (12.6) | 0.027 | -0.22 |
| Week 1 | 198 | 18.7 (15.1) | 189 | 15.3 (17.0) | 0.000 | 0.90 | | 229 | 25.4 (16.7) | 215 | 18.9 (18.4) | | 0.000 | 1.03 | -3.6 (17.7) | 0.043 | -0.20 |
| Week 2 | 189 | 4.6 (11.0) | 182 | 1.0 (14.2) | 0.342 | 0.07 | | 208 | 5.4 (11.2) | 196 | -0.5 (17.2) | | 0.706 | -0.03 | 1.5 (15.8) | 0.368 | 0.09 |
| Week 4 | 178 | 3.3 (11.2) | 170 | -0.4 (12.8) | 0.706 | -0.03 | | 195 | 3.1 (8.6) | 182 | -2.8 (15.6) | | 0.015 | -0.18 | 2.5 (14.3) | 0.108 | 0.17 |
| Hours worked | | | | | | | | | | | | | | | | | |
| Baseline^e^ | 192 | 34.4 (16.8) |  |  |  |  | | 215 | 33.8 (16.0) |  |  | |  |  | 0.6 (16.4) | 0.721 | 0.04 |
| Week 1 | 198 | 19.2 (16.3) | 189 | -14.4 (18.1) | 0.000 | -0.79 | | 228 | 15.2 (14.9) | 213 | -18.9 (20.6) | | 0.000 | -0.92 | 4.5 (19.5) | 0.020 | 0.23 |
| Week 2 | 189 | 31.6 (16.0) | 182 | -2.4 (16.4) | 0.049 | -0.15 | | 207 | 33.3 (13.6) | 196 | -1.4 (17.4) | | 0.245 | -0.08 | -1.0 (16.9) | 0.584 | -0.06 |
| Week 4 | 178 | 34.8 (13.6) | 170 | 0.9 (16.7) | 0.475 | 0.05 | | 195 | 35.7 (13.3) | 182 | 0.8 (16.8) | | 0.540 | 0.05 | 0.2 (16.7) | 0.933 | 0.01 |
| PROMIS Fatigue 8(a) | | | | | | | | | | | | | | | | | |
| Baseline^e^ | 283 | 44.6 (8.9) |  |  |  |  | | 281 | 45.1 (9.9) |  |  | |  |  | -0.5 (9.4) | 0.560 | -0.05 |
| Week 1 | 282 | 59.9 (9.6) | 280 | 15.5 (11.1) | 0.000 | 1.40 | | 277 | 60.5 (10.0) | 276 | 15.3 (11.6) | | 0.000 | 1.31 | 0.2 (11.4) | 0.849 | 0.02 |
| Week 2 | 269 | 50.9 (10.0) | 267 | 6.3 (10.3) | 0.000 | 0.61 | | 261 | 51.1 (10.0) | 260 | 6.2 (11.5) | | 0.000 | 0.53 | 0.1 (10.9) | 0.899 | 0.01 |
| Week 4 | 260 | 46.9 (10.1) | 258 | 2.3 (9.4) | 0.000 | 0.24 | | 246 | 48.1 (10.1) | 245 | 3.1 (10.9) | | 0.000 | 0.28 | -0.8 (10.2) | 0.386 | -0.08 |

^a^ Score ranges: EQ-5D-5L VAS 0 to 100, EQ-5D-5L UI (the United States weights) -0.573 to 1; WPAI-GH (absenteeism, presenteeism, work productivity loss, and activity impairment) 0 to 100 percent.

^b^ Baseline refers to pre-COVID-19 symptom onset.

^c^ P-value of t-test comparing mean score changes from baseline and 0 within BNT162b2 or Unvaccinated cohorts.

^d^ ES_w_ refers to the standardized effect size for score changes from baseline within BNT162b2 or Unvaccinated cohorts.

^e^ P-value of t-test comparing mean score changes from baseline between BNT162b2 and Unvaccinated cohorts.

^f^ ES_b_ refers to the standardized effect size for score changes from baseline between BNT162b2 and Unvaccinated cohorts.

Supplemental Figure 1 SARS-CoV-2 Variants proportions in the US

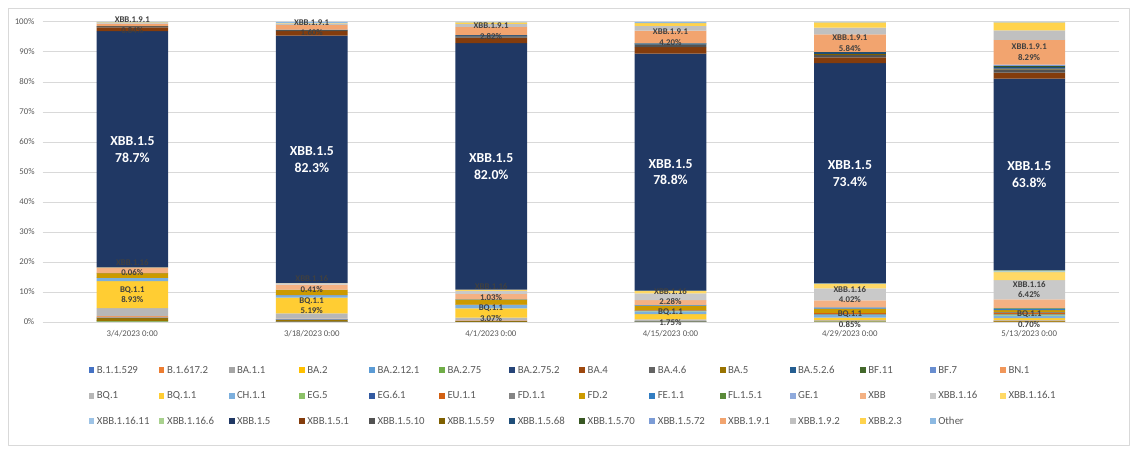

Source: reproduced and adapted from [SARS-CoV-2 Variant Proportions | Data | Centers for Disease Control and Prevention (cdc.gov)](https://data.cdc.gov/Laboratory-Surveillance/SARS-CoV-2-Variant-Proportions/jr58-6ysp).

An analysis of CDC variants data revealed that XBB.1.5 was predominant throughout the study period, ranging from 63.8% - 82.3%. BQ.1.1 was up to 8.9% in early March, and fell throughout April. XBB.1.16 and XBB.1.9.1 increased throughout April/May to 6-10% of cases.

Supplemental Figure 2 Prevalence of acute COVID-19 symptoms by vaccination status

(a) Time of testing

|  | Bivalent BNT162b2 | Unvaccinated |
| --- | --- | --- |
| Fever | 37.7% | 45.6% |
| Chills | 44.3% | 58.7% |
| Muscle or Body Aches | 39.6% | 52.6% |
| Headache | 60.4% | 69.1% |
| Fatigue | 63.9% | 67.9% |
| Shortness of Breath or Difficulty Breathing | 15.2% | 16.2% |
| Cough | 75% | 77.7% |
| Sore Throat | 60.4% | 60.9% |
| New/Recent Loss of Taste or Smell | 11.7% | 15.9% |
| Congestion or Runny Nose | 81.6% | 81.7% |
| Nausea or Vomiting | 3.16% | 4.59% |
| Diarrhea | 10.4% | 14.4% |

(b) Week 1

|  | Bivalent BNT162b2 | Unvaccinated |
| --- | --- | --- |
| Fever | 1.05% | 4.27% |
| Chills | 3.51% | 3.91% |
| Muscle or Body Aches | 11.9% | 18.9% |
| Headache | 17.5% | 28.1% |
| Fatigue | 48.8% | 52.3% |
| Shortness of Breath or Difficulty Breathing | 15.4% | 18.1% |
| Cough | 49.5% | 50.9% |
| Sore Throat | 13.7% | 14.2% |
| New/Recent Loss of Taste or Smell | 13% | 13.9% |
| Congestion or Runny Nose | 46.3% | 40.9% |
| Nausea or Vomiting | 0.35% | 0.71% |
| Diarrhea | 8.07% | 7.12% |

(c) Week 2

|  | Bivalent BNT162b2 | Unvaccinated |
| --- | --- | --- |
| Fever | 0.74% | 1.92% |
| Chills | 1.86% | 2.68% |
| Muscle or Body Aches | 13.4% | 16.9% |
| Headache | 12.3% | 17.6% |
| Fatigue | 40.1% | 42.1% |
| Shortness of Breath or Difficulty Breathing | 8.18% | 11.1% |
| Cough | 34.6% | 34.9% |
| Sore Throat | 6.32% | 10.3% |
| New/Recent Loss of Taste or Smell | 6.32% | 4.6% |
| Congestion or Runny Nose | 23.8% | 23.4% |
| Nausea or Vomiting | 0.74% | 0.38% |
| Diarrhea | 4.83% | 6.13% |

(d) Week 4

|  | Bivalent BNT162b2 | Unvaccinated |
| --- | --- | --- |
| Fever | 0.77% | 1.22% |
| Chills | 1.15% | 1.22% |
| Muscle or Body Aches | 6.54% | 10.2% |
| Headache | 11.5% | 15.1% |
| Fatigue | 17.7% | 31.4% |
| Shortness of Breath or Difficulty Breathing | 8.46% | 11.4% |
| Cough | 16.5% | 20.4% |
| Sore Throat | 2.31% | 6.12% |
| New/Recent Loss of Taste or Smell | 3.08% | 4.08% |
| Nausea or Vomiting | 2.69% | 4.49% |
| Diarrhea | 1.92% | 4.08% |
